# Exosome-Based Therapy for Spinal Cord Injury Repair: A Systematic Review of Preclinical Evidence and Exploratory Quantitative Synthesis

**DOI:** 10.64898/2026.07.27.26359044

**Authors:** Farzan Fahim, Hossein Mahmoodi, Amirmahdi Mojtahedzadeh, Mahsa Faramin Lashkarian, Mana Majlesi, Mojtaba Esmaeeli, Hossein Sattari, Mohammad Maroufi, Parmida Mafakhery, Seyed Younes Hashemi, Samin Koohi Kamali, Mahdieh Mansoori, Elaheh Aghazadeh, Saeed Safari, Fatemeh Khazaei, Alireza Zali

**Author notes:** Farzan Fahim and Hossein Mahmoodi contributed equally to this work and share first authorship. **Co-corresponding authors: Farzan Fahim,** Department of Neurosurgery, Shohada-e Tajrish Hospital, Shahid Beheshti University of Medical Sciences, Tehran, Iran, Men’s Health and Reproductive Health Research Center, Shahid Beheshti University of Medical Sciences, Tehran, Iran, **Email:**, **Saeed Safari,** Men’s Health and Reproductive Health Research Center, Shahid Beheshti University of Medical Sciences, Tehran, Iran, Emergency Care Promotion Research Center, Shahid Beheshti University of Medical Sciences, Tehran, Iran, **Email:**.

## Abstract

**Background:** Exosome- and extracellular vesicle-based therapies have emerged as promising cell-free approaches for spinal cord injury repair, with reported effects on inflammation, apoptosis, myelination, axonal regeneration, angiogenesis, blood–spinal cord barrier integrity, and neurogenesis. However, the preclinical evidence is heterogeneous, and the extent to which the available data permit quantitative synthesis remains unclear.

**Methods:** This systematic review was conducted in accordance with PRISMA 2020 and registered in PROSPERO as a preclinical animal intervention review (CRD420261446664). PubMed/MEDLINE, Scopus, Web of Science, and Embase were searched from inception to 1 June 2026 without language or publication-date restrictions. Eligible studies evaluated an exosome- or extracellular vesicle-based intervention for spinal cord injury and reported functional, histological, molecular, electrophysiological, vascular, regenerative, or safety-related outcomes. Risk of bias was assessed using an adapted version of SYRCLE’s tool for animal studies, while the first-in-human phase I study was appraised separately using the JBI Critical Appraisal Checklist for Quasi-Experimental Studies. Study characteristics, intervention strategies, outcome domains, and risk-of-bias patterns were synthesized descriptively. Where complete group-level means, standard deviations, and sample sizes were available, exploratory quantitative synthesis was performed using standardized mean differences calculated as Hedges’ *g*.

**Results:** The search identified 1,329 records. After removal of 481 duplicates, 848 records were screened, 108 full-text reports were assessed for eligibility, and 24 studies were included, comprising 23 animal/preclinical studies and one human phase I study. Exosome sources, injury models, administration routes, dosing strategies, and follow-up durations varied substantially. Reported outcomes included locomotor recovery, lesion and tissue preservation, myelination, axonal and neural regeneration, inflammation, apoptosis, angiogenesis, blood–spinal cord barrier repair, neurogenesis, and safety. Among the 23 animal studies, none was judged to be at overall low risk of bias; 20 were classified as unclear risk and three as high risk. The human phase I study was appraised separately and judged to be at high risk of bias for causal efficacy inference. Two studies contributed complete data to the exploratory meta-analysis of Basso, Beattie, and Bresnahan locomotor recovery. Both study-level estimates favored exosome treatment, while the random-effects pooled estimate was imprecise and crossed the null (Hedges’ *g* 3.74; 95% CI −0.53 to 8.00).

**Conclusions:** Exosome- and extracellular vesicle-based therapies demonstrated promising signals across functional and biological domains of spinal cord injury repair. However, the evidence was limited by methodological heterogeneity, unclear risk of bias, inconsistent reporting of vesicle characterization and dosing, and insufficient complete numerical data for robust quantitative synthesis. Preregistered, adequately powered, and transparently reported studies using standardized intervention and outcome-reporting methods are required to clarify therapeutic efficacy and translational potential.

## Introduction

Spinal cord injury (SCI) can cause persistent motor, sensory, and autonomic dysfunction, particularly after severe or complete injury. Although advances in acute management and rehabilitation have improved survival and supportive care, restoration of damaged neural tissue and meaningful neurological recovery remain limited. This unmet therapeutic need has stimulated increasing interest in regenerative and cell-free approaches designed to preserve tissue, modulate secondary injury, and promote neural repair.

The biological response to SCI involves multiple interacting pathological processes. Neuroinflammation and dysregulated immune-cell activity contribute to secondary tissue injury, as demonstrated by studies targeting macrophage polarization, microglial responses, neutrophil extracellular-trap formation, and inflammatory cytokine signaling [1,3,8,16,19]. Oxidative stress and apoptosis also contribute to neuronal loss, with several experimental interventions acting through Bax/Bcl-2 signaling, oxidative-stress pathways, and other regulators of cell survival [3,10,18,22]. Additional therapeutic targets investigated in the available literature include astrocytic activation [5], blood–spinal cord barrier dysfunction [15], vascular repair and angiogenesis [14,23], axonal regeneration [7,12,17], and endogenous neurogenesis [24]. Because these processes overlap, interventions capable of influencing several biological pathways simultaneously may be particularly relevant to SCI repair.

Exosomes and extracellular vesicles have emerged as promising cell-free therapeutic platforms because they can carry biologically active molecules and can be modified to influence specific cellular pathways. Investigated strategies have included miRNA-loaded vesicles derived from induced pluripotent stem cells [2], miR-21-containing mesenchymal stromal cell exosomes [10], circBDNF-engineered exosomes targeting PI3K/AKT/mTOR signaling [7], miR-29b-3p-containing extracellular vesicles targeting the PTEN/Akt/mTOR pathway [22], and PTEN-siRNA-loaded exosomes administered intranasally [9]. Other approaches have incorporated pharmacological cargo [3,5,8], bioactive peptides [1,17], hypoxic preconditioning [18], and biomaterial-based delivery systems intended to improve local retention and sustained release [6,11–13,20,21].

The experimental landscape is highly diverse. Vesicles have been derived from umbilical cord mesenchymal stromal/stem cells [4,12,15,22], placental mesenchymal stromal cells [23,24], macrophages [8], plasma [3], induced pluripotent stem cell-derived cells [2], urine-derived stem cells [6], cortical neurons [21], and other mesenchymal stromal cell populations [9,11,14,16,18,19]. Administration routes have included intravenous delivery [16,18,19,22], intrathecal injection [4], intranasal administration [9], and local delivery at or near the lesion site [6,11–13,20,21]. Reported outcomes have included locomotor recovery, tissue preservation, neural and axonal regeneration, inflammation, apoptosis, angiogenesis, blood–spinal cord barrier repair, neurogenesis, autonomic recovery, and safety [1–24].

Despite these encouraging findings, interpretation of the evidence remains difficult because studies vary substantially in species, SCI model and severity, vesicle source, isolation and characterization methods, cargo modification, dose definition, administration route, treatment timing, follow-up duration, and outcome measurement. Incomplete numerical reporting further limits direct comparison and quantitative synthesis. Therefore, this systematic review aimed to synthesize the available evidence on exosome- and extracellular vesicle-based therapies for SCI, characterize heterogeneity in experimental and intervention-related features, assess methodological risk of bias, and perform an exploratory quantitative synthesis of locomotor recovery where sufficiently complete data were available.

## Methods

### Protocol and reporting framework

This systematic review was conducted according to a protocol specifying the review question, eligibility criteria, search strategy, study-selection process, data-extraction framework, risk-of-bias assessment, and synthesis methods. The protocol was registered in PROSPERO as a preclinical animal intervention review under registration number CRD420261446664 and is publicly available through the PROSPERO database [26]. The registration process was informed by PROSPERO4animals guidance [27].

The review was reported in accordance with the Preferred Reporting Items for Systematic Reviews and Meta-Analyses 2020 statement [25]. The completed PRISMA 2020 checklist is provided as Supplementary file S4.

### Search strategy and information sources

PubMed/MEDLINE, Scopus, Web of Science, and Embase were searched from database inception to 1 June 2026. No restrictions were applied according to publication date or language. Potentially eligible non-English reports were translated where necessary and assessed using the same eligibility criteria as English-language reports.

The search strategy combined controlled-vocabulary and free-text terms relating to three principal concepts: spinal cord injury, exosomes or extracellular vesicles, and functional or neurological recovery. Database-specific syntax was adapted to the indexing requirements of each information source. The complete search strategies for all databases, including the search terms, Boolean operators, and any limits applied, are provided in Supplementary file S1.

### Eligibility criteria

Studies were eligible if they investigated an exosome- or extracellular vesicle-based intervention in the context of spinal cord injury and reported at least one relevant functional, histological, molecular, imaging, electrophysiological, inflammatory, apoptotic, regenerative, vascular, blood–spinal cord barrier, autonomic, or safety-related outcome.

Eligible interventions included native exosomes or small extracellular vesicles, engineered or cargo-modified vesicles, exosome-loaded biomaterials, hydrogel- or scaffold-based delivery systems, and extracellular vesicles derived from stem or stromal cells, plasma, immune cells, neural cells, or other biologically relevant sources. Preclinical animal intervention studies constituted the principal evidence base. Clinically oriented or quasi-experimental studies were also eligible when they directly evaluated exosome- or extracellular vesicle-based therapy for spinal cord injury; these studies were synthesized and appraised separately from the animal evidence.

Reviews, editorials, commentaries, conference abstracts without extractable primary data, retracted articles, studies unrelated to spinal cord injury, studies without an exosome or extracellular vesicle intervention, and studies that did not report relevant recovery, repair, or safety outcomes were excluded. The excluded full-text reports and corresponding reasons for exclusion are provided in Supplementary file S3.

### Study selection

All retrieved records were imported into EndNote 2025 and deduplicated before screening. Two reviewers independently screened titles and abstracts against the predefined eligibility criteria. Records considered potentially eligible by either reviewer were advanced to full-text assessment.

Full-text reports were independently assessed by two reviewers using the same eligibility framework. Disagreements were resolved through discussion and consensus, with unresolved decisions adjudicated by a third reviewer. The study-selection process was documented using a PRISMA 2020 flow diagram.

### Data extraction and outcomes

Data were extracted using a standardized form developed for this review. Two reviewers independently performed data extraction, and all entries were verified against the original reports. Discrepancies were resolved by consensus, with final adjudication by a third reviewer where required. The complete extraction dataset is provided as Supplementary file S2.

Extracted variables included bibliographic information, study design, animal species and strain, sex, age, body weight, sample size, spinal cord injury model, injury mechanism and severity, injury level, experimental groups, vesicle-producing cell source, tissue and species of origin, culture conditions, isolation and purification methods, characterization methods, particle size and concentration, cargo or surface engineering, dose, dose unit, administration route, treatment timing, dosing frequency, follow-up duration, biodistribution or targeting information, functional outcomes, imaging findings, histological outcomes, molecular outcomes, inflammatory and apoptotic markers, angiogenic and blood–spinal cord barrier markers, neurogenesis-related outcomes, electrophysiological outcomes, autonomic outcomes, safety findings, and risk-of-bias information.

The primary outcome was locomotor functional recovery at the latest eligible follow-up, assessed principally using the Basso, Beattie, and Bresnahan locomotor rating scale in rat studies and the Basso Mouse Scale in mouse studies. Secondary outcomes included lesion or cavity size and tissue preservation; neuronal survival and axonal regeneration; myelination and oligodendrocyte preservation; glial scar and astrocytic responses; inflammatory and immune markers; apoptosis and other cell-death outcomes; angiogenesis and blood–spinal cord barrier repair; neurogenesis; electrophysiological recovery; autonomic outcomes; and safety.

### Risk-of-bias assessment

Risk of bias in animal intervention studies was assessed using an adapted version of SYRCLE’s risk-of-bias tool [28]. The assessed domains comprised sequence generation, baseline similarity, random housing, blinding of caregivers or investigators, random outcome assessment, blinding of outcome assessors, incomplete outcome data, selective outcome reporting, and other potential sources of bias. Each domain was categorized as low, high, or unclear risk of bias, or as not applicable where appropriate.

Domain-level judgments constituted the primary risk-of-bias assessment. Overall study-level classifications were used descriptively. A study was categorized as high risk when at least one assessed domain was judged high risk, as unclear risk when no domain was judged high risk but at least one domain was unclear, and as low risk when all applicable domains were judged low risk. Overall classifications were not treated as weighted numerical scores.

The first-in-human phase I study was unsuitable for assessment using the adapted SYRCLE framework and was appraised separately using the JBI Critical Appraisal Checklist for Quasi-Experimental Studies [29]. Item-level judgments and the overall appraisal are presented in Table 6. All risk-of-bias assessments were performed independently by two reviewers, with disagreements resolved through consensus or adjudication by a third reviewer.

### Data synthesis

A structured descriptive synthesis was conducted for all eligible studies. Study and intervention characteristics were organized according to population or species, spinal cord injury model, injury mechanism, vesicle source, intervention type, engineering strategy, administration route, treatment timing, dosing approach, follow-up duration, and reported outcome domains.

Descriptive subgroup summaries were prepared according to species, spinal cord injury model, vesicle source, intervention type, and administration route. These summaries were presented using study counts and proportions. Outcome-domain availability was mapped across functional, histological, molecular, inflammatory, apoptotic, myelination, axonal-regeneration, vascular, blood–spinal cord barrier, neurogenesis, electrophysiological, autonomic, and safety outcomes.

### Quantitative analysis

Studies were eligible for quantitative synthesis when sufficiently comparable outcomes were reported with complete group-level means, standard deviations, and sample sizes for both the exosome-based intervention and spinal cord injury control groups. The latest eligible follow-up was selected for the primary locomotor analysis. When studies reported multiple eligible intervention groups or time points, comparisons were selected according to the prespecified review question, and shared control groups were not counted more than once within the same synthesis.

Continuous outcomes were expressed as standardized mean differences calculated using Hedges’ *g*, which applies a small-sample correction to Cohen’s *d*. Positive effect estimates indicated outcomes favoring exosome- or extracellular vesicle-based treatment. For outcomes in which lower values represented improvement, the direction of effect was reversed before analysis to ensure consistent interpretation.

Random-effects models were fitted using restricted maximum likelihood estimation because biological and methodological heterogeneity was anticipated across animal models, injury paradigms, vesicle sources, doses, administration routes, treatment timing, and follow-up periods. Summary effects were reported as Hedges’ *g* with 95% confidence intervals. Statistical heterogeneity was assessed using Cochran’s Q, the I² statistic, and the estimated between-study variance, τ².

Forest plots were used to display individual and pooled effect estimates. Secondary outcomes with complete numerical data but insufficiently comparable or statistically dependent observations were displayed as individual standardized effects without pooled estimates. Funnel plots and statistical tests for small-study effects were planned only for syntheses containing at least 10 independent studies. Meta-regression, formal subgroup meta-analysis, and leave-one-out sensitivity analysis were planned only when the number and comparability of available studies permitted meaningful interpretation.

All statistical analyses and visualizations were conducted using R version 4.6.0. Effect-size calculations and meta-analytic models were implemented using the **metafor** package [30]. Data management and visualization were performed using **readxl, dplyr, tidyr, janitor, writexl, and ggplot2**.

## Results

### Study selection

The database search identified 1,329 records before deduplication, including 313 from PubMed/MEDLINE, 316 from Scopus, 394 from Web of Science, and 306 from Embase. After removal of 481 duplicate records, 848 records underwent title and abstract screening. Of these, 740 were excluded because they were clearly irrelevant (n = 420), unrelated to spinal cord injury (n = 185), or unrelated to exosomes or extracellular vesicles (n = 135).

A total of 108 reports were sought for retrieval, all of which were successfully obtained and assessed at full text. Eighty-four reports were excluded: review article (n = 62), not related to spinal cord injury (n = 11), no exosome or extracellular vesicle intervention (n = 6), neither exosome-related nor spinal cord injury-related (n = 3), editorial article (n = 1), and retracted article (n = 1).

Twenty-four reports met the eligibility criteria and were included in the descriptive synthesis [1–24]. These comprised 23 animal/preclinical studies and one first-in-human phase I study. Two studies provided complete numerical data suitable for the exploratory quantitative synthesis of Basso, Beattie, and Bresnahan locomotor recovery [9,11]. The study-selection process is presented in the PRISMA 2020 flow diagram in Figure 1, and the excluded full-text reports with corresponding reasons are listed in Supplementary file S3.

**Figure 1.**
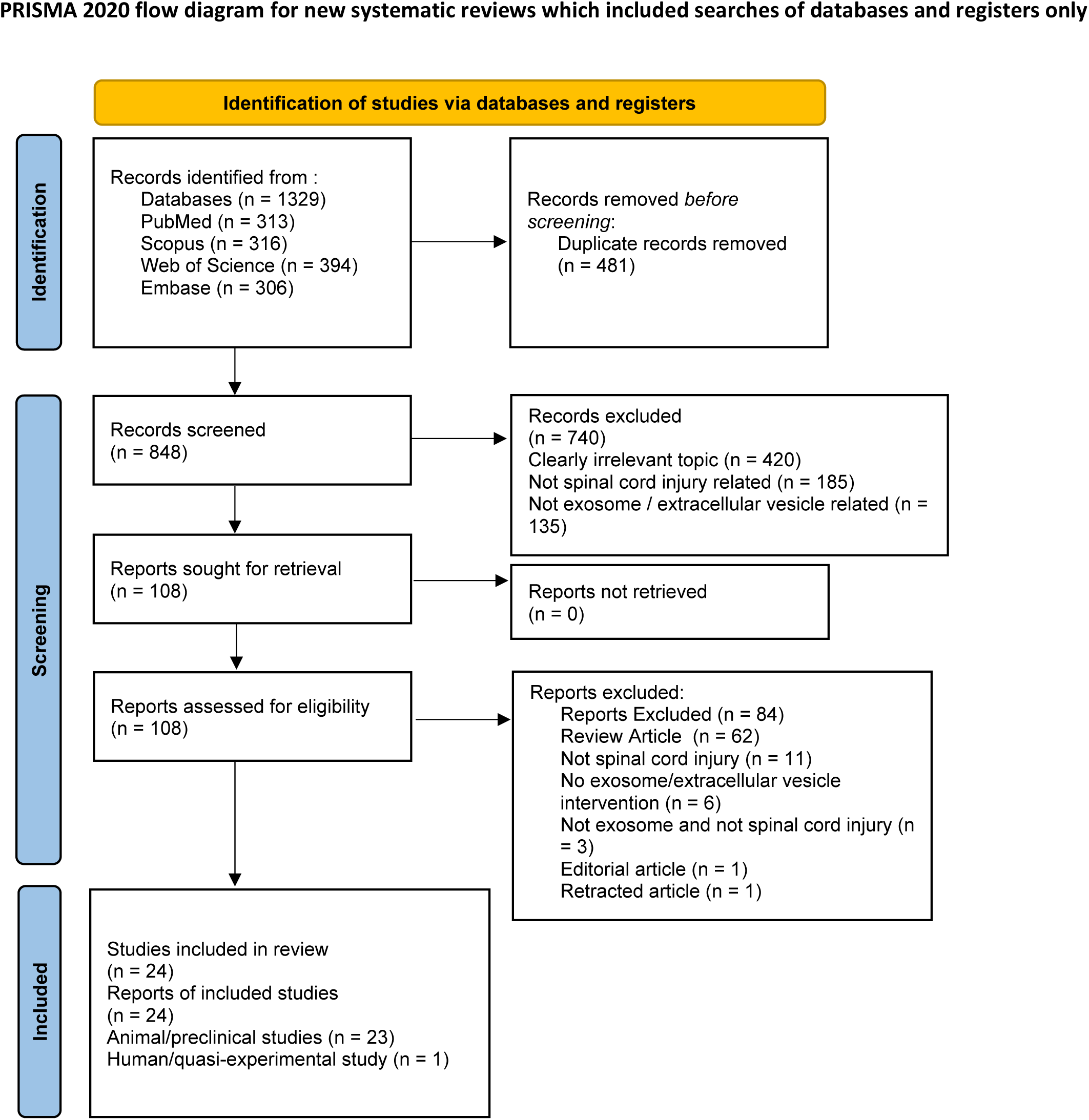
PRISMA 2020 flow diagram of study selection. Flow diagram summarizing record identification, duplicate removal, title/abstract screening, full-text eligibility assessment, exclusions with reasons, and final inclusion. The search identified 1,329 records from PubMed/MEDLINE, Scopus, Web of Science, and Embase. After removal of 481 duplicate records, 848 records underwent title and abstract screening, of which 740 were excluded. A total of 108 reports were assessed for eligibility, and 84 were excluded with documented reasons. Twenty-four reports were included in the review, comprising 23 animal/preclinical studies and one first-in-human phase I study. Two studies contributed complete extractable data to the exploratory quantitative synthesis of BBB locomotor recovery.

### Characteristics of included studies

The 24 included reports were published between 2018 and 2026 and evaluated exosome- or extracellular vesicle-based interventions for spinal cord injury repair [1–24]. Twelve studies used rat models, 11 used mouse models, and one was a human phase I study. Contusion or impact injury was the most frequently used experimental paradigm (n = 12), followed by transection or hemisection (n = 7), compression or clip injury (n = 4), and another or clinical injury context (n = 1). Injury sites were predominantly thoracic. The distributions of study population or species and injury model are presented in Figure 2A and Figure 2B.

**Figure 2.**
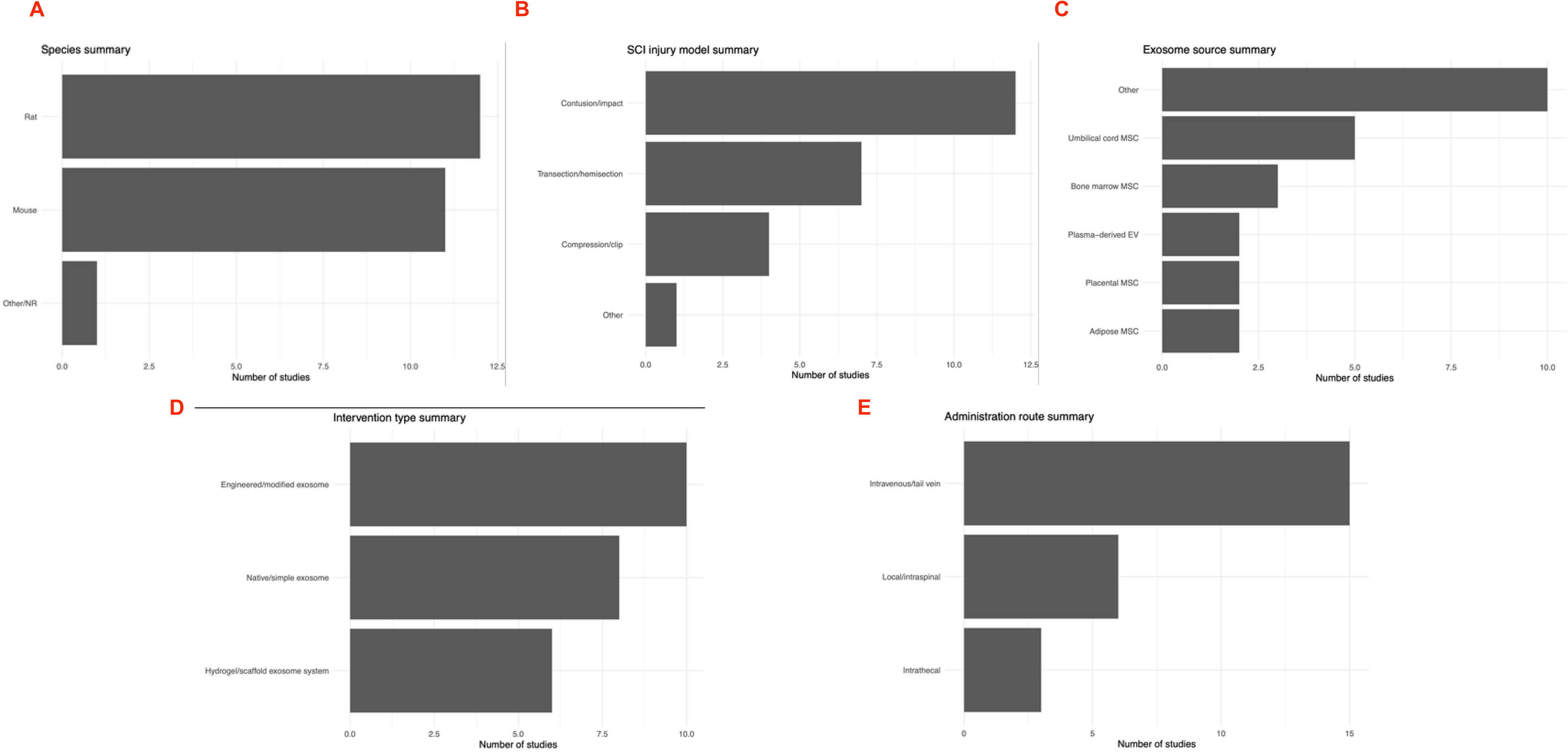
Descriptive characteristics of included studies. Multi-panel summary of study and intervention characteristics across the 24 included reports. (A) Distribution according to study population or species, including rat, mouse, and human/other. (B) Distribution according to spinal cord injury model or context, including contusion/impact, transection/hemisection, compression/clip, and other or clinical injury contexts. (C) Distribution according to vesicle source, categorized as umbilical cord MSC, bone marrow MSC, adipose MSC, placental MSC, plasma-derived extracellular vesicles, or other sources. (D) Distribution according to intervention strategy, categorized as engineered or modified vesicles, native or minimally modified vesicles, or hydrogel/scaffold-associated delivery systems. (E) Distribution according to administration route, including intravenous/tail-vein, local/intraspinal, and intrathecal delivery. Together, these panels demonstrate substantial heterogeneity in experimental models, vesicle sources, therapeutic engineering strategies, and delivery approaches.

Vesicle sources varied substantially. Umbilical cord mesenchymal stromal/stem cells were represented in five studies, bone marrow mesenchymal stromal/stem cells in three, and adipose-derived, placental, and plasma-derived vesicles in two studies each. The remaining studies used other cellular sources or engineered producer systems. The distribution of vesicle sources is presented in Figure 2C.

Intervention strategies included engineered or cargo-modified vesicles (n = 10), native or minimally modified vesicles (n = 8), and hydrogel- or scaffold-associated vesicle systems (n = 6). Intravenous or tail-vein administration was the predominant delivery category, followed by local or intraspinal and intrathecal administration. Considerable variation was observed in treatment timing, administration frequency, follow-up duration, and dose reporting. Doses were expressed using particle number, protein concentration, injection volume, or incompletely standardized descriptions, preventing meaningful dose harmonization or dose–response analysis. Intervention types and administration routes are summarized in Figure 2D and Figure 2E.

The main characteristics of the included studies are presented in Table 1, and the complete extraction dataset is provided in Supplementary file S2.

**Table 1.** Characteristics of included studies. Summary of the 24 included reports evaluating exosome- or extracellular vesicle-based interventions for spinal cord injury, including 23 animal/preclinical studies and one first-in-human phase I study. The table includes study identification, publication year, country, study type, animal or participant characteristics where applicable, spinal cord injury model or clinical injury context, intervention source, delivery route, timing, follow-up duration, and main reported outcomes. The full extraction dataset is provided in Supplementary file S2.

### Outcome domains

Locomotor functional recovery was the principal outcome domain and was assessed primarily using the Basso, Beattie, and Bresnahan locomotor rating scale in rat studies and the Basso Mouse Scale in mouse studies. BBB outcomes were reported more frequently than BMS outcomes.

Myelination was the most frequently reported outcome domain (n = 22). Lesion, cavity, or tissue-preservation outcomes and axonal or neural regeneration were each reported in 16 studies. BBB locomotor scores were reported in 15 studies, inflammatory outcomes in 14, apoptosis-related outcomes in 11, and BMS scores in eight.

Additional outcomes included neuronal survival, oligodendrocyte preservation, glial and astrocytic responses, angiogenesis, blood–spinal cord barrier integrity, neurogenesis, electrophysiological recovery, autonomic function, and safety [1–24]. Outcome-domain availability indicates whether a domain was reported and does not necessarily indicate that complete numerical data were available for quantitative synthesis. Most secondary outcomes were therefore synthesized descriptively. The distribution of outcome availability is presented in Figure 3.

**Figure 3.**
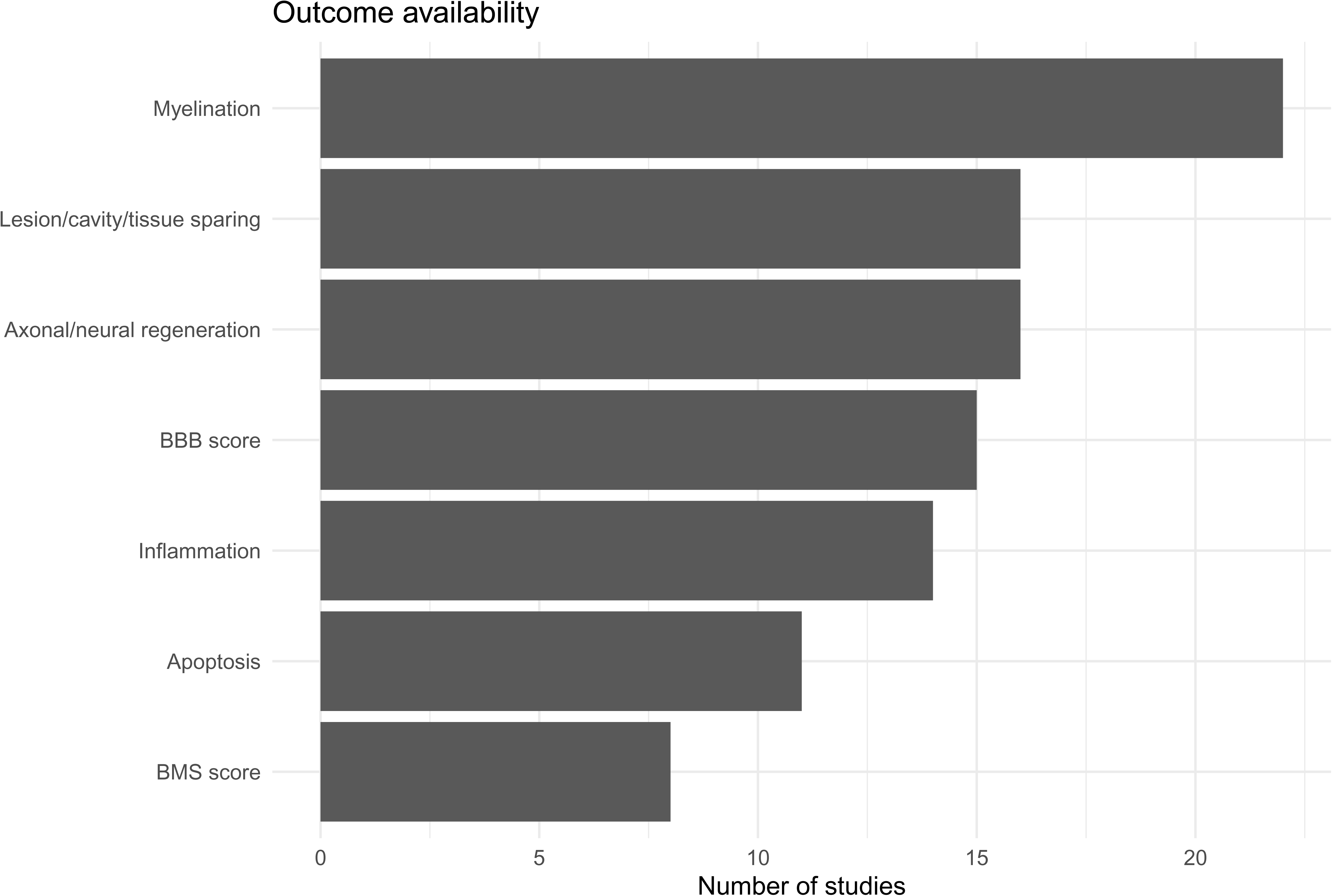
Outcome-domain availability across included studies. Bar plot showing the number of studies reporting each major outcome domain. Outcomes included functional recovery measures such as BBB and BMS scores, as well as biological and mechanistic domains including lesion/cavity/tissue preservation, inflammation, apoptosis, myelination, and axonal/neural regeneration. Availability reflects whether an outcome domain was reported and does not necessarily indicate availability of complete numerical data for meta-analysis.

### Risk-of-bias assessment

The 23 animal intervention studies were assessed using the adapted SYRCLE risk-of-bias framework [28], while the first-in-human phase I study was appraised separately using the JBI Critical Appraisal Checklist for Quasi-Experimental Studies. The adapted SYRCLE judgments are presented in Table 2 and Figure 4, and the item-level JBI appraisal is presented in Table 6.

**Figure 4.**
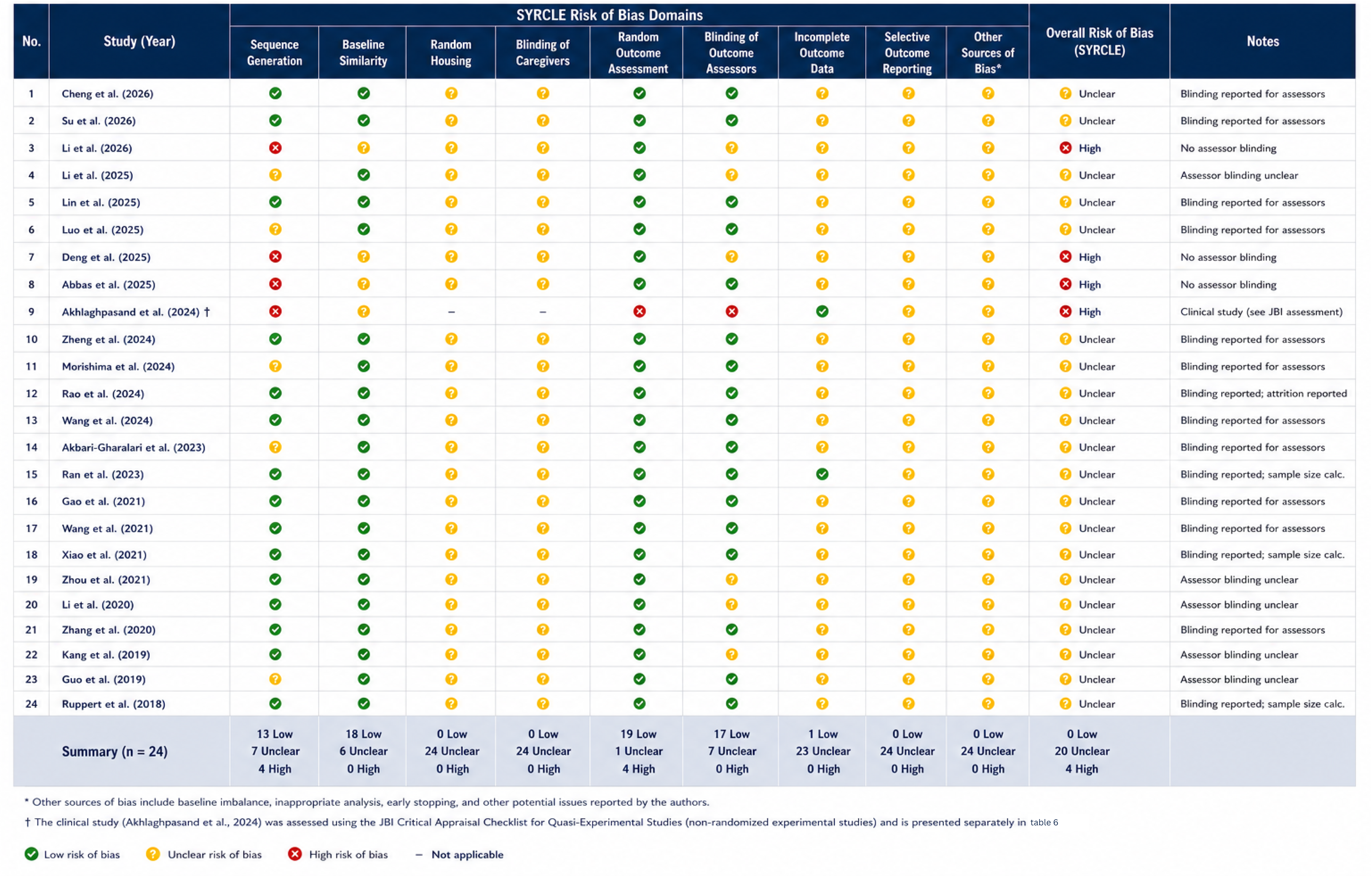
SYRCLE risk-of-bias summary. Study-level judgments across the adapted SYRCLE risk-of-bias domains for the 23 included animal intervention studies. The assessed domains were sequence generation, baseline similarity, random housing, blinding of caregivers or investigators, random outcome assessment, blinding of outcome assessors, incomplete outcome data, selective outcome reporting, and other potential sources of bias. The first-in-human phase I study is annotated for completeness but was excluded from the SYRCLE summary and appraised separately using the JBI Critical Appraisal Checklist for Quasi-Experimental Studies; its item-level appraisal is presented in Table 6. Green indicates low risk of bias, yellow indicates unclear risk, red indicates high risk, and a dash indicates not applicable.

**Table 2.** Risk-of-bias assessment of included studies. Study-level judgments for the 23 animal intervention studies using an adapted version of SYRCLE’s risk-of-bias tool. The assessed domains comprised sequence generation, baseline similarity, random housing, blinding of caregivers or investigators, random outcome assessment, blinding of outcome assessors, incomplete outcome data, selective outcome reporting, and other potential sources of bias. Domain-level judgments constituted the primary assessment. Overall classifications were used descriptively and were not treated as weighted numerical scores. The first-in-human phase I study was assessed separately using the JBI Critical Appraisal Checklist for Quasi-Experimental Studies and is presented in Table 6.

**Table 3.** Exosome source, engineering strategy, dose, route, and timing. Summary of intervention characteristics across included studies, including exosome or extracellular vesicle source, parent-cell origin, isolation method, engineering or cargo modification, biomaterial incorporation, dose reporting, administration route, number of injections, treatment timing after injury, and follow-up duration. This table highlights heterogeneity in exosome production, dosing, and delivery strategies.

**Table 4.** Outcome domains and synthesis approach. Summary of primary and secondary outcomes reported across included studies and the corresponding synthesis method used in this review. Locomotor recovery was considered the primary outcome and was assessed using BBB or BMS scores. Secondary domains included lesion/cavity/tissue preservation, neuronal survival and axonal regeneration, myelination, glial scar response, inflammation, apoptosis, angiogenesis and blood–spinal cord barrier repair, neurogenesis, electrophysiology, and safety. Domains with complete numerical data were considered for quantitative synthesis; others were synthesized descriptively.

**Table 5.** Exploratory quantitative synthesis of BBB locomotor recovery. Study-level and pooled standardized mean differences for BBB locomotor recovery in studies with complete extractable mean, standard deviation, and sample-size data. Positive Hedges’ g values indicate greater locomotor recovery in exosome-treated animals compared with spinal cord injury controls. The pooled estimate was generated using a random-effects model and should be interpreted as exploratory because only two studies contributed data.

**Table 6.** JBI critical appraisal of the included human phase I study. Item-level appraisal of Akhlaghpasand et al. [4] using the JBI Critical Appraisal Checklist for Quasi-Experimental Studies. Q1: Was the cause-and-effect relationship clearly established in temporal sequence? Q2: Were the participants included in the comparisons similar? Q3: Did the participants receive similar treatment or care apart from the intervention of interest? Q4: Was there an independent control group? Q5: Were there multiple measurements of the outcome before and after the intervention? Q6: Was follow-up complete, and were differences between participants completing and not completing follow-up adequately described and analyzed? Q7: Were outcomes measured consistently across the compared time points? Q8: Were outcomes measured reliably? Q9: Was appropriate statistical analysis used? The study was retained because it provided directly relevant first-in-human evidence regarding the safety and feasibility of intrathecal exosome administration. The absence of an independent control group and multiple pre-intervention measurements substantially limited causal interpretation of efficacy outcomes. Because the JBI checklist does not prescribe a weighted numerical summary score, the overall judgment was assigned descriptively and applies specifically to causal efficacy inference rather than to the relevance of the reported safety findings.

Among the animal studies, none was classified as overall low risk of bias; 20 were categorized as overall unclear risk and three as high risk. The human phase I study was retained as directly relevant safety and feasibility evidence but was judged to be at high risk of bias for causal efficacy inference. It satisfied criteria relating to temporal sequence, participant comparability, consistency of concurrent care, completeness of follow-up, and consistent outcome measurement. However, it lacked an independent control group and multiple pre-intervention measurements, while the reliability of outcome assessment and adequacy of the statistical analysis for causal efficacy inference were unclear.

Unclear judgments predominated for random housing, blinding of caregivers or investigators, incomplete outcome data, selective outcome reporting, and other potential sources of bias. Sequence generation showed a mixture of low-, unclear-, and high-risk judgments. Baseline similarity and random outcome assessment were more frequently judged to be at low risk, while blinding of outcome assessors was reported more favorably than most other domains.

### Exploratory quantitative synthesis of BBB locomotor recovery

Two studies reported complete treatment- and control-group means, standard deviations, and sample sizes for BBB locomotor recovery and were included in the exploratory quantitative synthesis [9,11]. Outcomes were analyzed at the latest eligible follow-up using standardized mean differences calculated as Hedges’ *g*, with positive estimates favoring exosome-based treatment.

Both study-level estimates favored treatment. Guo et al. reported a large effect on BBB locomotor recovery (Hedges’ *g* 6.01; 95% CI 3.86 to 8.16) [9], whereas Li et al. reported a smaller positive effect (Hedges’ *g* 1.66; 95% CI 0.48 to 2.83) [11].

The random-effects pooled estimate favored exosome-based treatment but was imprecise and crossed the null (Hedges’ *g* 3.74; 95% CI −0.53 to 8.00). Statistical heterogeneity was substantial (Q = 12.11, df = 1, p < 0.001; I² = 91.7%; τ² = 8.68), indicating marked variation in effect magnitude between the two studies. The pooled estimate should therefore be interpreted as exploratory rather than as a precise estimate of treatment efficacy.

The forest plot is presented in Figure 5, and the individual study-level estimates are additionally displayed in Supplementary Figure S1. Because only two independent studies contributed to the synthesis, funnel-plot asymmetry assessment, statistical testing for small-study effects, meta-regression, formal subgroup meta-analysis, and leave-one-out sensitivity analysis were not undertaken.

**Figure 5.**
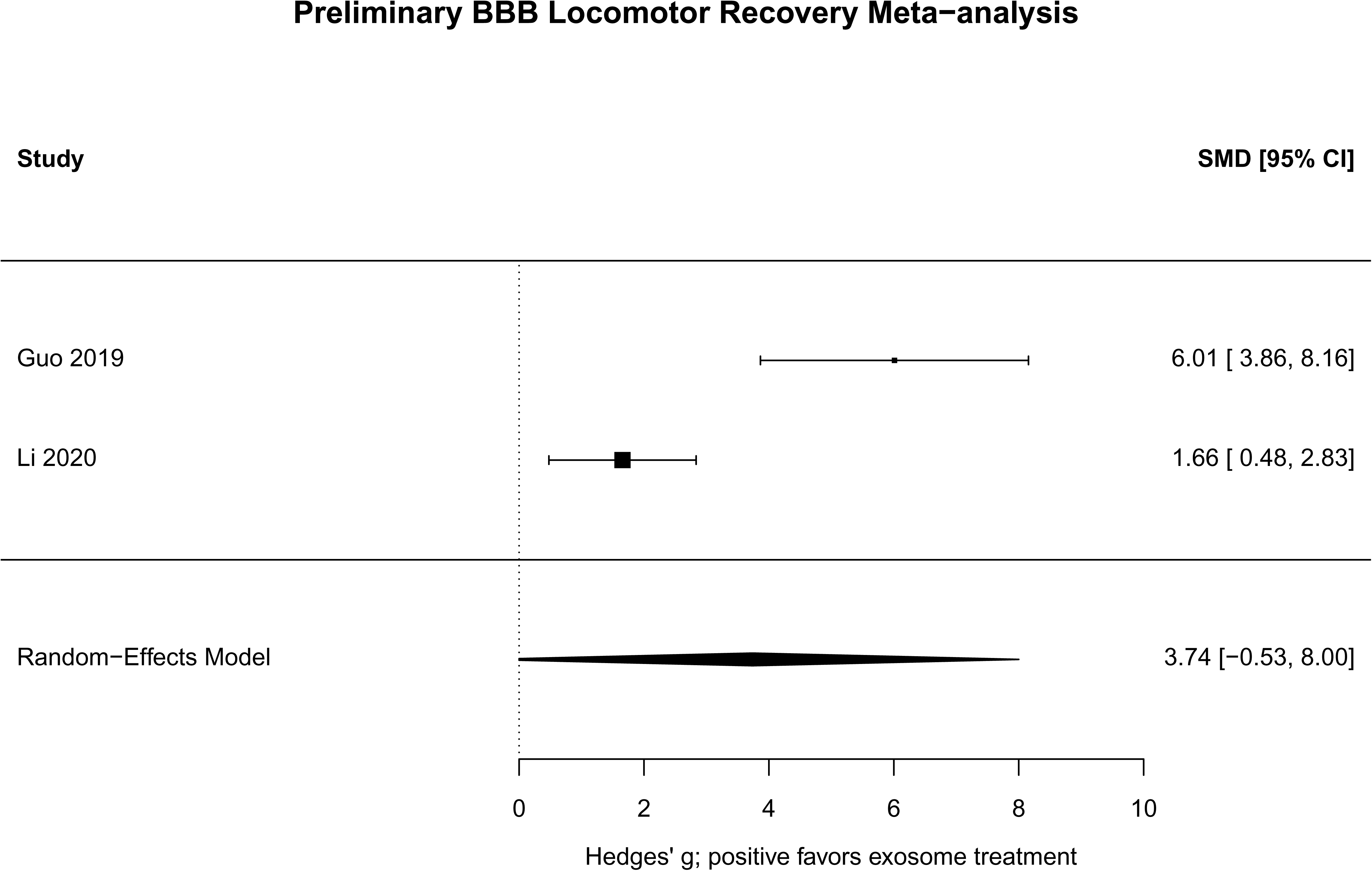
Exploratory meta-analysis of BBB locomotor recovery. Random-effects forest plot of standardized mean differences for BBB locomotor recovery among studies with complete extractable mean, standard deviation, and sample-size data. Positive Hedges’ g values favor exosome-based treatment over spinal cord injury control. Because only two studies were eligible for quantitative synthesis, this analysis was considered exploratory and was not used for funnel plot asymmetry testing, meta-regression, formal subgroup analysis, or leave-one-out sensitivity analysis.

### Secondary quantitative effects

Complete numerical data were available for a limited number of secondary outcomes. These included caudal cross-sectional tissue area and motor-evoked-potential amplitude reported by Guo et al. [9], together with several neurogenesis-related cellular markers reported by Zhou et al. [24].

The displayed effect estimates favored exosome-based treatment, but no pooled estimate was calculated because the outcomes represented isolated comparisons or multiple statistically dependent markers derived from the same study. These findings should therefore be interpreted as exploratory and hypothesis-generating rather than as independent pooled evidence. The secondary effect-size plot is presented in Supplementary Figure S2.

### Descriptive subgroup findings

Formal subgroup meta-analysis was not performed because the quantitative evidence base was insufficient for reliable inferential comparisons. Study distributions were instead summarized descriptively according to species, spinal cord injury model, vesicle source, intervention type, and administration route.

Rat and mouse models were both well represented, contusion or impact injury was the predominant experimental paradigm, intravenous or tail-vein administration was the most frequent delivery category, and engineered or cargo-modified vesicles constituted the largest intervention category. These descriptive distributions are presented in Figure 2.

Overall, the included studies reported generally favorable functional and biological signals across diverse experimental settings. However, variation in injury model, vesicle source, engineering strategy, dose definition, administration route, treatment timing, follow-up duration, and outcome reporting substantially limited direct comparison and quantitative inference.

## Discussion

### Main findings

This systematic review synthesized 24 reports evaluating exosome- and extracellular vesicle-based interventions for spinal cord injury (SCI), comprising 23 animal/preclinical studies and one first-in-human phase I study [1–24]. Across the included evidence, vesicle-based interventions were generally associated with favorable signals in locomotor recovery, tissue preservation, myelination, axonal and neural regeneration, neuroinflammation, apoptosis, angiogenesis, blood–spinal cord barrier integrity, and neurogenesis. However, substantial variation in experimental models, vesicle sources, engineering strategies, doses, administration routes, treatment timing, follow-up durations, and outcome measurements limited direct comparison across studies.

Locomotor recovery was the most clinically relevant functional outcome and was evaluated predominantly using the Basso, Beattie, and Bresnahan locomotor rating scale in rats and the Basso Mouse Scale in mice. Complete numerical data suitable for quantitative synthesis were available for only two BBB studies [9,11]. Both individual estimates favored exosome-based treatment, but the random-effects pooled estimate was imprecise and crossed the null (Hedges’ *g* 3.74; 95% CI −0.53 to 8.00). Statistical heterogeneity was substantial (I² = 91.7%), indicating marked variation in effect magnitude between the two studies. The pooled estimate should therefore be interpreted as exploratory and hypothesis-generating rather than as a definitive measure of treatment efficacy.

The principal contribution of this review lies in mapping the diversity of the experimental and therapeutic landscape, identifying recurring biological pathways, assessing methodological limitations, and demonstrating the extent to which incomplete numerical reporting restricts robust meta-analysis.

### Biological mechanisms of exosome-mediated repair

The therapeutic rationale for exosome- and extracellular vesicle-based SCI treatment is supported by the multifactorial nature of secondary spinal cord injury. Rather than acting through a single pathway, the included interventions appeared to influence inflammatory activity, oxidative stress, apoptosis, vascular integrity, tissue preservation, myelination, and regenerative signaling.

Immunomodulation was among the most frequently investigated mechanisms. Several studies reported reductions in pro-inflammatory cytokine signaling, altered microglial or macrophage activation, suppression of neutrophil extracellular-trap formation, or promotion of reparative immune phenotypes [1,3,8,16,19,20]. These findings suggest that vesicle-based treatments may attenuate components of the inflammatory microenvironment that contribute to continuing neural and glial injury.

Anti-apoptotic and neuroprotective effects were also frequently described. Experimental interventions were associated with modulation of Bax/Bcl-2 signaling, reductions in apoptotic or oxidative-stress markers, and improvements in neuronal-survival indicators [3,10,18,22]. Other engineered preparations acted through signaling pathways related to neural survival and regeneration, including PI3K/AKT/mTOR and PTEN/Akt/mTOR signaling [7,22]. These findings support the possibility that vesicle cargo may reduce secondary cell death and preserve vulnerable neural tissue surrounding the lesion.

Favorable changes in myelination, axonal regeneration, and neural plasticity were reported using markers such as MBP, NF200, MAP2, TUJ1, GAP43, NeuN, and synaptophysin, together with histological measures of tissue preservation and neural organization [6,7,9,11–13,18,20–24]. These outcomes are biologically relevant because functional recovery depends not only on limiting lesion expansion but also on preserving axonal conduction, supporting oligodendrocytes, and restoring neural connectivity.

Vascular and blood–spinal cord barrier repair represented another important mechanistic domain. Some studies reported increased angiogenic signaling, improved endothelial or tight-junction integrity, and reduced barrier permeability following treatment [14,15,23]. Such effects may reduce edema and inflammatory-cell infiltration while improving the local environment for tissue repair.

A substantial proportion of the evidence involved engineered or biomaterial-associated vesicle systems. Investigated approaches included miRNA- or siRNA-loaded vesicles [2,9,10,22], circRNA-engineered exosomes [7], pharmacologically loaded vesicles [3,5,8], peptide-modified preparations [1,17], hypoxia-preconditioned vesicles [18], and hydrogel- or scaffold-based delivery platforms [6,11–13,20,21]. These strategies may enhance cargo specificity, lesion targeting, or local retention, but they also introduce additional sources of heterogeneity and make it difficult to distinguish the independent therapeutic effect of the vesicles from that of the incorporated cargo or biomaterial.

### Interpretation of the evidence

The direction of the included evidence was generally favorable, but the strength of inference remained limited. Most studies reported improvement in at least one functional, structural, or molecular outcome [1–24]. Nevertheless, positive findings from individual experiments should be interpreted cautiously because differences in design, injury severity, intervention composition, comparator selection, dosing, and follow-up can substantially influence observed treatment effects.

The risk-of-bias assessment further reduced confidence in the evidence. Among the 23 animal studies, none was classified as overall low risk; 20 were categorized as unclear risk and three as high risk. Unclear judgments predominated particularly for random housing, blinding of caregivers or investigators, incomplete outcome data, selective outcome reporting, and other potential sources of bias. The first-in-human phase I study was appraised separately and judged to be at high risk of bias for causal efficacy inference because it lacked an independent control group and multiple pre-intervention measurements. These judgments do not necessarily invalidate the reported findings, but they limit confidence in internal validity and causal interpretation.

The present review therefore provides a more cautious interpretation than would be obtained by considering the favorable findings of individual studies in isolation. The available evidence supports continued investigation of exosome- and extracellular vesicle-based therapy, but it does not yet establish a precise, reproducible, or clinically transferable magnitude of benefit.

### Clinical and translational implications

Exosome- and extracellular vesicle-based interventions offer a flexible cell-free platform that can be engineered, loaded with therapeutic cargo, and combined with biomaterials. The included studies demonstrate the feasibility of systemic, intrathecal, intranasal, and local delivery, as well as the potential use of sustained-release hydrogel or scaffold systems [4,6,9,11–13,16,18–22]. The first-in-human phase I study also provided preliminary information regarding the feasibility and safety of intrathecal administration in patients with complete subacute SCI [4]. However, evidence of clinical efficacy remains insufficient.

A major barrier to translation is the absence of standardized vesicle production and characterization. The included studies varied in parent-cell source, culture conditions, isolation and purification methods, characterization markers, particle-size reporting, particle concentration, protein concentration, storage procedures, and dose units. These differences impair reproducibility and make cross-study comparisons difficult.

Dose and route optimization also remain unresolved. Systemic administration may be less invasive and more clinically practical, but it may lead to off-target biodistribution and reduced lesion-site exposure. Local or intraspinal administration may improve target delivery but is more invasive. Biomaterial-supported delivery may prolong local retention and release, although the additional scaffold or hydrogel components increase manufacturing and regulatory complexity [6,11–13,20,21].

Treatment timing is another critical consideration. Acute, subacute, and chronic SCI represent biologically distinct environments, and an intervention that attenuates early inflammation may not necessarily promote axonal or circuit reconstruction during later phases. Future studies should therefore define injury phase and treatment timing clearly and should avoid combining biologically distinct time windows without justification.

Outcome selection should also become more clinically relevant and standardized. BBB and BMS scores remain important measures of locomotor recovery, but future translational studies should also assess sensory function, autonomic recovery, electrophysiology, imaging, tissue preservation, biodistribution, and long-term safety. Development of a core outcome set for preclinical SCI vesicle studies would substantially improve comparability.

### Future directions

Future preclinical investigations should use adequately justified sample sizes, clearly described randomization procedures, blinded outcome assessment, random housing, transparent eligibility and attrition criteria, and prespecified statistical analyses. These measures would reduce unclear risk-of-bias judgments and improve confidence in the resulting evidence.

Complete numerical reporting is essential. Studies should provide group means, standard deviations or standard errors, sample sizes, exact assessment time points, group definitions, and complete information regarding exclusions and missing observations. Frequent graphical reporting without accompanying numerical data was a major reason that most outcomes could not be quantitatively synthesized in this review.

Head-to-head studies are needed to determine whether therapeutic effects vary according to vesicle source, engineering strategy, dose, administration route, treatment timing, or injury model. Direct comparisons of native versus engineered vesicles, systemic versus local delivery, single versus repeated administration, and free versus biomaterial-retained vesicles would be particularly informative.

Mechanistic investigations should move beyond isolated marker changes. Integrated transcriptomic, proteomic, lipidomic, and spatial analyses could help identify the vesicle components and recipient-cell pathways most closely associated with functional recovery. Such approaches may distinguish general trophic effects from pathway-specific regenerative mechanisms.

Translational research should also prioritize biodistribution, immunogenicity, off-target organ accumulation, dose-related toxicity, manufacturing scalability, batch consistency, storage stability, and long-term safety. These domains were inconsistently addressed in the current evidence base.

### Limitations

This review has several limitations. First, although 24 studies were included, formal quantitative synthesis was restricted to two BBB studies because most outcomes lacked complete extractable means, standard deviations, and sample sizes [9,11]. The pooled estimate was therefore highly uncertain and should not be interpreted as a definitive measure of efficacy.

Second, substantial biological and methodological heterogeneity existed across species, injury models, lesion severity, vesicle sources, isolation and characterization methods, doses, administration routes, treatment timing, and follow-up duration. This heterogeneity limited pooling and precluded reliable formal subgroup analysis.

Third, risk-of-bias assessment identified frequent unclear reporting, particularly for random housing, caregiver or investigator blinding, incomplete outcome data, selective outcome reporting, and other potential sources of bias. These limitations reduce confidence in the internal validity and reproducibility of the evidence.

Fourth, publication bias and small-study effects could not be assessed because the quantitative synthesis contained too few independent studies. Unpublished negative findings, preprints, and non-indexed studies may therefore have been missed despite the absence of language or publication-date restrictions.

Fifth, several secondary outcomes were represented by isolated comparisons or multiple correlated markers from the same study, preventing independent pooled analysis. These findings should be regarded as exploratory.

Finally, a formal GRADE assessment was not undertaken because the evidence base was predominantly preclinical and the quantitative synthesis was exploratory.

## Conclusion

Exosome- and extracellular vesicle-based interventions were generally associated with favorable preclinical signals across locomotor recovery, tissue preservation, myelination, axonal regeneration, inflammation, apoptosis, angiogenesis, blood–spinal cord barrier integrity, and neurogenesis. However, the evidence remains limited by substantial methodological heterogeneity, incomplete reporting, unclear risk of bias, inconsistent vesicle characterization and dose definition, and insufficient complete numerical data for robust meta-analysis.

The exploratory synthesis of BBB locomotor recovery favored treatment, but its wide confidence interval and substantial heterogeneity preclude definitive conclusions regarding effect magnitude. Preregistered, adequately powered, transparently reported, and methodologically standardized studies are required before exosome- and extracellular vesicle-based therapies can be advanced confidently toward clinical application.

## Supporting information

Tabels

## Data Availability

All data analyzed in this review were extracted from published studies. The complete data extraction sheet, excluded-study table, search strategies, risk-of-bias assessment, PRISMA checklist, and supplementary analyses are provided as supplementary materials. The statistical code used for descriptive analyses and exploratory quantitative synthesis can be made available from the corresponding author upon reasonable request.

## Declarations

### Ethics approval and consent to participate

Ethics approval was not required for this systematic review because the study synthesized data from previously published articles and did not involve direct experimentation on humans or animals by the authors. For the included primary studies, ethics approval and animal-care statements were extracted where reported.

### Use of artificial intelligence tools

AI-assisted tools were used to support language editing, formatting, and organization of the manuscript. All scientific content, data extraction, interpretation, and final wording were reviewed, verified, and approved by the authors. No AI tool was used as a substitute for independent scientific judgment.

## Supplementary material legends

**Supplementary file S1. Complete database search strategies.**

Full search syntax used for PubMed/MEDLINE, Scopus, Web of Science, and Embase. Searches combined terms for spinal cord injury, exosomes/extracellular vesicles, and functional or neurological recovery. Searches were performed from database inception to 1 June 2026 without language or date restrictions.

**Supplementary file S2. Full data extraction sheet.**

Complete extraction table for all 24 included studies. Extracted variables include study identification, experimental design, animal characteristics, spinal cord injury model, exosome source and characterization, dose and delivery details, functional outcomes, histological outcomes, molecular outcomes, safety outcomes, and risk-of-bias fields.

**Supplementary file S3. Excluded studies with reasons.**

List of excluded articles with documented reasons for exclusion. Exclusion categories included review article, not spinal cord injury, not exosome/extracellular vesicle intervention, neither exosome nor spinal cord injury, editorial article, and retracted article.

**Supplementary file S4. PRISMA 2020 checklist.**

Completed PRISMA 2020 checklist indicating where each reporting item is addressed in the manuscript and supplementary materials.

**Supplementary Figure S1. Study-level effect sizes for BBB locomotor recovery.**

Plot of individual standardized mean differences for BBB recovery in the two studies with complete extractable numerical data. Positive values favor exosome-based treatment. This figure complements the exploratory BBB forest plot.

**Supplementary Figure S2. Secondary extractable single-study effects.**

Forest-style plot of standardized effects for secondary outcomes with complete numerical data. No pooled estimate was calculated because these outcomes were either single-study comparisons or multiple biological markers derived from the same study.

## Abbreviations

BBB: Basso, Beattie, and Bresnahan locomotor rating scale
Bcl-2: B-cell lymphoma 2
BMS: Basso Mouse Scale
CI: Confidence interval
EV: Extracellular vesicle
GRADE: Grading of Recommendations Assessment, Development and Evaluation
JBI: Joanna Briggs Institute
MAP2: Microtubule-associated protein 2
MBP: Myelin basic protein
MEP: Motor evoked potential
miRNA / miR: MicroRNA
MSC: Mesenchymal stromal/stem cell
NF200: Neurofilament 200
Nrf2: Nuclear factor erythroid 2-related factor 2
PRISMA: Preferred Reporting Items for Systematic Reviews and Meta-Analyses
PTEN: Phosphatase and tensin homolog
Q: appraisal question
ROS: Reactive oxygen species
SCI: Spinal cord injury
SD: Standard deviation
SE: Standard error
SIRT1: Sirtuin 1
SYRCLE: Systematic Review Centre for Laboratory Animal Experimentation
TNF-α: Tumor necrosis factor alpha
Treg: Regulatory T cell
TUJ1: Neuron-specific class III beta-tubulin
VEGF: Vascular endothelial growth factor

