## Supplementary figures and images for "Exosome-Based Therapy for Spinal Cord Injury Repair: A Systematic Review of Preclinical Evidence and Exploratory Quantitative Synthesis"

### Supplementary Figure S1

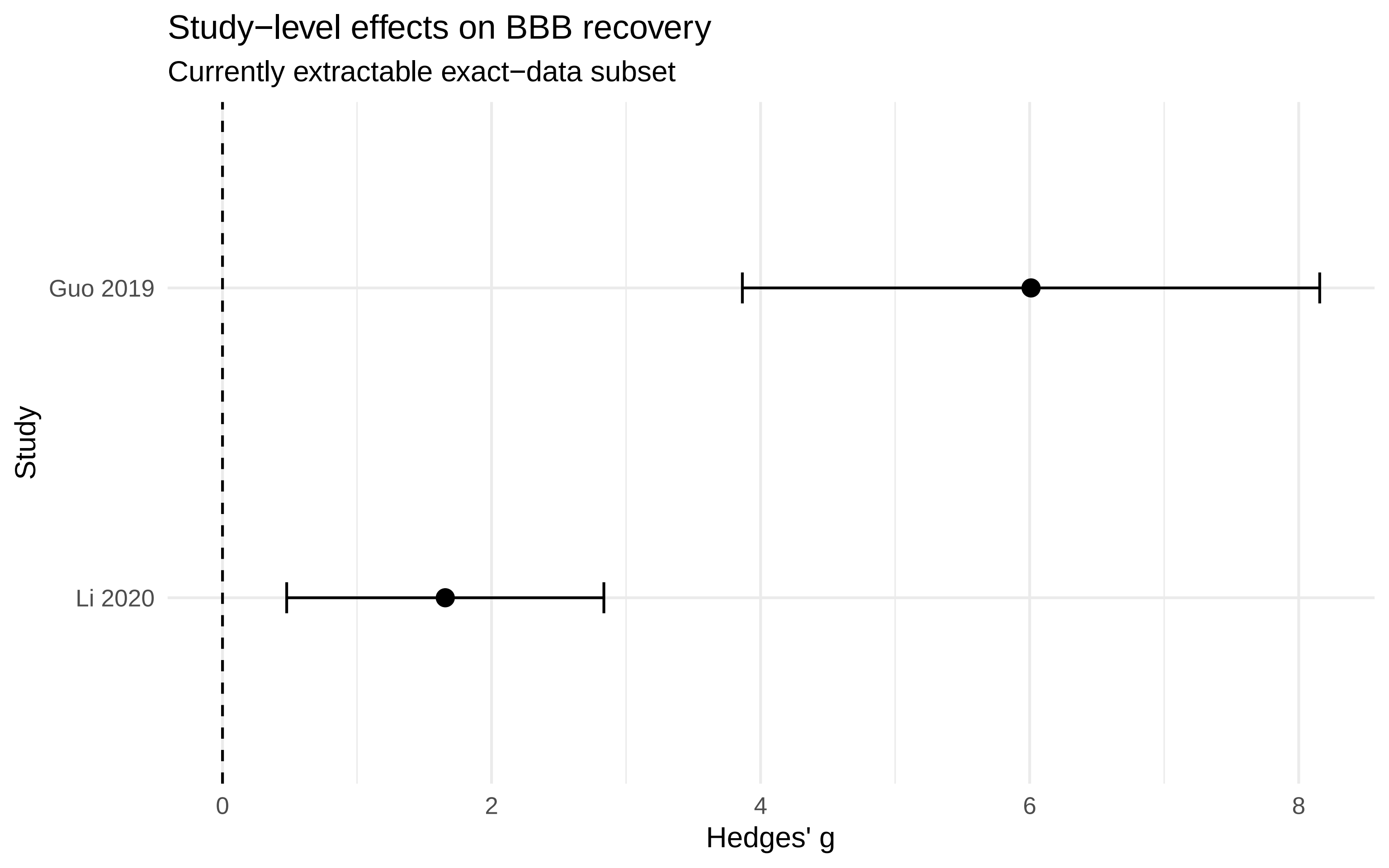

### Supplementary Figure S2

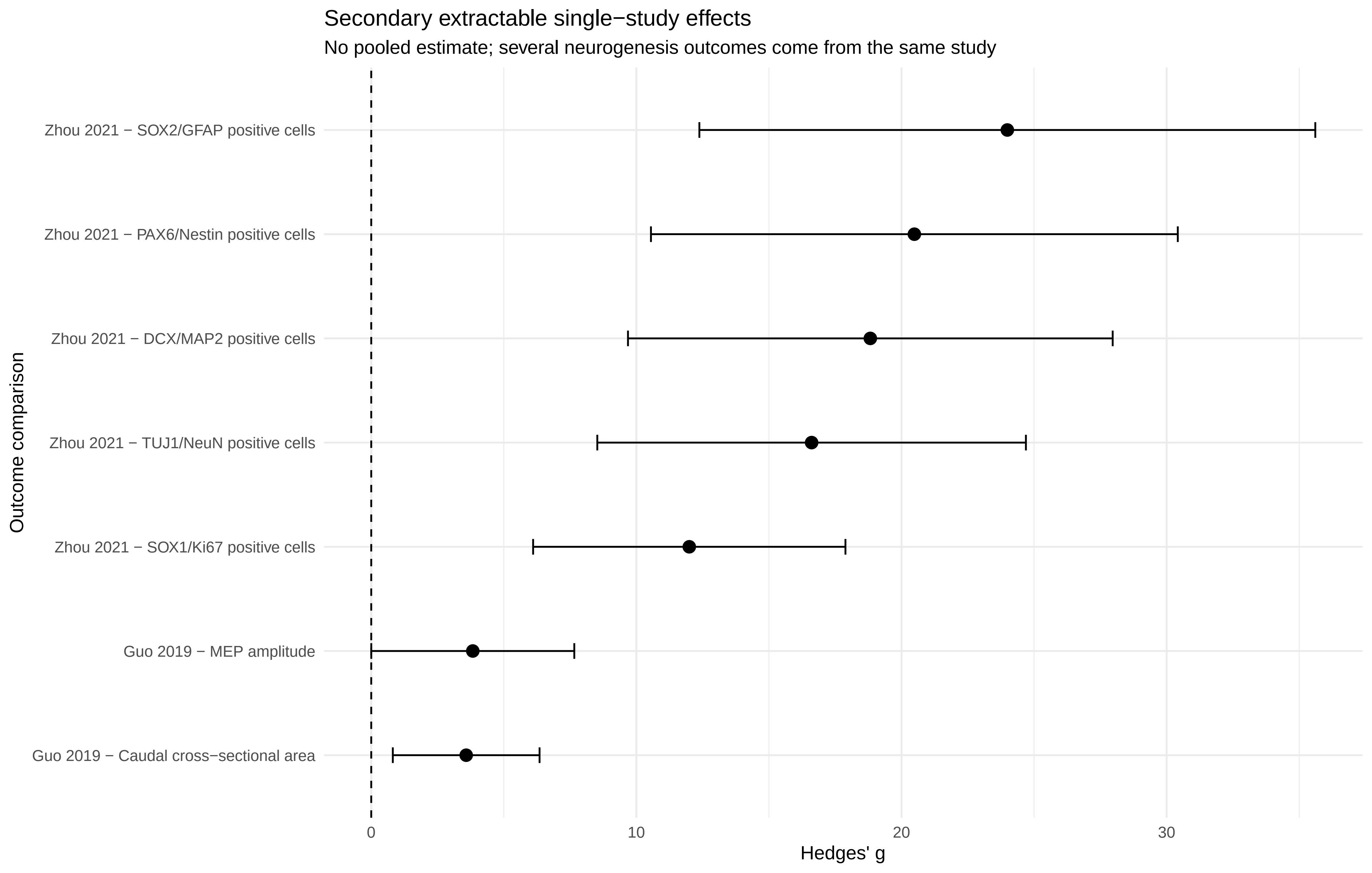
