## Supplementary file S1 for "Exosome-Based Therapy for Spinal Cord Injury Repair: A Systematic Review of Preclinical Evidence and Exploratory Quantitative Synthesis"

Pubmed ;313

("spinal cord injury" OR "spinalcord injuries" OR "spinal injury" OR "SCI" OR"acute spinal cord injury" OR "chronic spinal cord injury"OR "traumatic spinal cord injury" OR "spinal trauma") AND(exosome* OR "extracellular vesicle*" OR EVs OR "small extracellularvesicle*" OR "cell-derived vesicle*" OR "secretedvesicle*" OR "exosome therapy" OR "exosome treatment"OR "exosomal therapy" OR "exosome-based" OR "MSCexosome*" OR "stem cell exosome*" OR "mesenchymal stem cellexosome*") AND ("functional recovery" OR "motorrecovery" OR "sensory recovery" OR "neurologicalrecovery" OR "motor function" OR "functional outcome*"OR "locomotor recovery" OR "locomotion" OR "behavioralrecovery" OR "Basso" OR "BBB score" OR "BassoBeattie Bresnahan" OR "ASIA score" OR "ASIA impairmentscale")

*________________________________*

scopus =316

TITLE-ABS-KEY(

("spinal cord injury" OR "spinal cordinjuries" OR "spinal injury" OR SCI OR "acute spinal cordinjury" OR

"chronic spinal cord injury" OR "traumaticspinal cord injury" OR "spinal trauma")AND

(ex

osome* OR "extracellular vesicle*" OR EVs OR"small extracellular vesicle*" OR "cell-derived vesicle*" OR "

secreted vesicle*" OR "exosometherapy" OR "exosome treatment" OR "exosomal therapy" OR "e

xosome-based" OR "MSC exosome*" OR"stem cell exosome*" OR "mesenchymal stem cell exosome*")AND("f

unc

tional recovery" OR "motorrecovery" OR "sensory recovery" OR "neurologicalrecovery" OR "moto

r function" OR "functionaloutcome*" OR "locomotor recovery" OR locomotion OR "behav

ioral recovery" OR Basso OR "BBBscore" OR "Basso Beattie Bresnahan" OR "ASIA s

core" OR "ASIA impairmentscale"))

*____________________________________________*

wos-394

TS=(

("spinal cord injury" OR "spinal cordinjuries" OR "spinal injury" OR "SCI" O

R "acute spinal cord injury" OR "chronicspinal cord injury" OR

"traumatic spinal cord injury" OR "spinaltrauma")AND

(ex

osome* OR "extracellular vesicle*" OR EVs OR"small extracellular vesicle*" OR "

cell-derived vesicle*" OR "secretedvesicle*" OR "exosome therapy" OR "e

xosome treatment" OR "exosomaltherapy" OR "exosome-based" OR "MS

C exosome*" OR "stem cellexosome*" OR "mesenchymal stem cell exosome*")AND("fu

nct

ional recovery" OR "motorrecovery" OR "sensory recovery" OR "neur

ological recovery" OR "motorfunction" OR "functional outcome*" OR "locom

otor recovery" OR locomotion OR"behavioral recovery" OR "Basso"

OR "BBB score" OR "BassoBeattie Bresnahan" OR "ASIA sc

ore" OR "ASIA impairmentscale"))

*_________________________________*

Embase =306

(

'spinal cordinjury'/exp

OR "spinalcord injury":ti,ab,kw

OR "spinalcord injuries":ti,ab,kw

OR "spinalinjury":ti,ab,kw

OR"SCI":ti,ab,kw	O

R "acutespinal cord injury":ti,ab,kw	OR

"chronicspinal cord injury":ti,ab,kw	OR

"traumaticspinal cord injury":ti,ab,kw	OR "

spinaltrauma":ti,ab,kw)AND(

'

exo

s

ome'/exp	OR 'e

xtracellular vesicle'/exp	ORexo

some*:ti,ab,kw	OR"ext

racellular vesicle*":ti,ab,kw	OR"EVs"

:ti,ab,kw	OR "smal

lextracellular vesicle*":ti,ab,kw	OR"cell-d

erived vesicle*":ti,ab,kw	OR "secret

edvesicle*":ti,ab,kw	OR "exosome

therapy":ti,ab,kw	OR "exosomet

reatment":ti,ab,kw	OR "exosomalt

herapy":ti,ab,kw	OR"exosome-bas

ed":ti,ab,kw	OR "MSCexosome*

":ti,ab,kw	OR "stem cellexo

some*":ti,ab,kw	OR"mesenchymal st

em cell exosome*":ti,ab,kw)AND(	'functionalre

c

ove

r

y'/exp	OR 'motorperformanc

e'/exp	OR 'sensoryfunction'

/exp	OR 'neurologicaldefic

it'/exp	OR"functional recovery

":ti,ab,kw	OR "motorrecovery":ti,a

b,kw	OR "sensoryrecovery":ti,

ab,kw	OR"neurological recovery"

:ti,ab,kw	OR "motorfunction":ti,ab,k

w	OR"functional outcome*":ti,

ab,kw	OR "locomotorrecovery":ti,ab

,kw	ORlocomotion:ti,ab,kw	OR"

behavioral recovery":ti,ab,

kw	OR Basso:ti,ab,kw	OR "BBB s

core":ti,ab,kw	OR "

BassoBeattie Bresnahan":ti,a

b,kw	OR "ASIAscore":ti,ab,kw	OR "A

SIAimpairment scale":ti,ab,kw

)
