## Supplementary material for "Exosome-Based Therapy for Spinal Cord Injury Repair: A Systematic Review of Preclinical Evidence and Exploratory Quantitative Synthesis": Tabels

**Table legends**

**Table 1. Characteristics of included studies.**

| Study | Country | Design / population | SCI model / injury context | Exosome or EV intervention | Delivery and timing | Follow-up | Main reported outcomes |
| --- | --- | --- | --- | --- | --- | --- | --- |
| Su Xinjin 2026 | China | Preclinical in vitro and in vivo mouse study | Female C57BL/6 mice; T10 contusive SCI | Engineered regulatory T-cell-derived exosomes with IKVAV surface modification | Tail-vein intravenous injection; single dose immediately after injury | 28 days | BMS recovery, Louisville Swim Scale, footprint parameters, bladder outcomes, inflammation, neuronal markers, myelination, axonal/neural regeneration, biodistribution and safety |
| Abbas 2024 | China | Preclinical in vitro and in vivo mouse study | BALB/c mice; weight-drop contusive SCI at T11–T12 | Bioengineered iPSC-derived exosomes with P2X3 targeting and miRNA loading | Intravenous administration; exact therapeutic timing not reported | 28 days | BMS recovery, neuronal survival, neurite outgrowth, histological morphology, apoptotic/inflammatory markers, biodistribution and systemic safety |
| Akbari-Gharalari 2023 | Iran | Preclinical in vivo mouse study | Female BALB/c mice; T10 compression SCI | Platelet-rich plasma-derived exosomes loaded with dexamethasone | Tail-vein intravenous injection; treatment started 1 h after SCI; repeated dosing | 28 days | BBB recovery, lesion area, inflammatory infiltration, cavity size, serum TNF-α/IL-10, Bax/Bcl-2 pathway markers |
| Akhlaghpasand 2024 | Iran | First-in-human single-arm open-label phase I study | Patients with complete subacute traumatic SCI; cervical or thoracic injury | Allogeneic human umbilical cord MSC-derived exosomes | Intrathecal injection via lumbar puncture; single dose 1–5.5 months after injury | 12 months | Safety, ASIA motor and sensory scores, SCIM III, bowel function, urinary function, spasticity and MRI safety assessment |
| Cheng 2026 | China | Preclinical in vitro and in vivo mouse study | Female C57BL/6 mice; T10 contusive SCI | hUCMSC-derived exosomes combined with ROS-responsive trifluoperazine prodrug nanoparticles | Systemic treatment after SCI; hUCMSC-exosomes once weekly for three injections | 6 weeks | BMS recovery, stride length, open-field activity, astrocyte phenotype, apoptosis, angiogenesis, neuronal preservation, cavity volume and tissue repair |
| Deng 2026 | China | Preclinical in vitro and in vivo mouse study | Female C57BL/6J mice; complete T10 transection | CD133+CD271+ human urine-derived stem cell exosomes incorporated into photosensitive hydrogel | Local injection into lesion site with hydrogel photocrosslinking immediately after transection | 56 days | BMS recovery, MEP recovery, bladder outcomes, angiogenesis, BSCB integrity, edema, Evans blue leakage, tight-junction proteins, axonal regeneration and neural repair |
| Li 2026 | China | Preclinical in vitro and in vivo mouse study | Male C57BL/6 mice; T10 clamp/compression SCI | HEK293T-derived exosomes engineered to carry circBDNF | Intrathecal injection at injury site immediately after SCI and on days 1 and 3 | 16 weeks | BMS recovery, gait parameters, MEP recovery, axonal regeneration, oxidative stress, apoptosis, inflammation, PI3K/AKT/mTOR signaling and neuroprotection |
| Gao 2021 | China | Preclinical in vitro and in vivo mouse study | C57BL/6J mice; Allen weight-drop contusive SCI at T9–T10 | M2 macrophage-derived exosomes loaded with berberine | Tail-vein intravenous injection; first dose 2 h after SCI; daily for 7 days | 28 days | BMS recovery, body weight, neuronal survival, inflammatory markers, macrophage polarization, apoptosis markers, biodistribution and pharmacokinetics |
| Guo 2019 | Israel / China | Preclinical in vitro and in vivo rat study | Female Sprague-Dawley rats; complete T10 transection | Human bone marrow MSC-derived exosomes loaded with PTEN siRNA | Intranasal repeated dosing beginning 2–3 h after SCI for 5 days; intralesional comparator | 8 weeks | BBB recovery, sensory recovery, urinary reflex, MRI/DTI metrics, MEP amplitude, axonal regeneration, angiogenesis, glial response and PTEN suppression |
| Kang 2019 | China | Preclinical in vitro and in vivo rat study | Sprague-Dawley rats; Allen contusive SCI at T9/T10 | MSC-derived exosomes enriched with miR-21 or PTEN-siRNA-related cargo | Intravenous injection after SCI; exact timing not reported | 28 days | BBB recovery, neuronal survival, apoptosis, miR-21/PTEN/PDCD4 signaling, caspase-3 expression and in vitro cell viability |
| Li 2020 | China | Preclinical biomaterial/exosome in vitro and in vivo rat study | Rats; long-span spinal cord transection | hMSC-derived exosomes immobilized in peptide-modified adhesive hydrogel scaffold | Local implantation into spinal cord lesion gap immediately after transection | 28 days | BBB recovery, ladder-rung performance, urinary outcomes, tissue regeneration, axonal regeneration, angiogenesis, glial scar response and exosome retention |
| Li 2025 | China | Preclinical in vitro and in vivo rat study | Rats; complete spinal cord transection defect model | Umbilical cord MSC-derived exosomes combined with coaxially printed NSC-laden microfibers | Local transplantation into complete spinal cord defect during SCI surgery | 4–8 weeks | BBB recovery, swimming/open-field outcomes, tissue bridging, neural regeneration, inflammation, apoptosis, scaffold integration and repair of large transection defects |
| Lin 2025 | China | Preclinical biomaterial/exosome in vitro and in vivo rat study | Female Sprague-Dawley rats; large-span complete transection at T9–T11 | Adipose MSC-derived exosomes embedded in ROS-responsive injectable hydrogel | Local injection into transection site immediately after injury | 6 weeks | BBB recovery, MEP recovery, bladder function, lesion repair, distal nerve repair, inflammation, apoptosis, myelination and axonal regeneration |
| Luo 2026 | China | Preclinical in vitro and in vivo rat study | Male Sprague-Dawley rats; T9–T10 weight-drop contusive SCI | Bone marrow MSC-derived exosomes combined with sodium tanshinone IIA sulfonate | Tail-vein exosome injection immediately after surgery plus oral pharmacological treatment | 21 days | BBB recovery, gait outcomes, lesion area, BSCB integrity, Evans blue leakage, VEGF signaling, inflammation, apoptosis and endothelial repair |
| Zheng 2024 | China | Preclinical in vivo rat study | Male Sprague-Dawley rats; modified Allen contusive SCI at T10 | Human umbilical cord MSC-derived exosomes | Tail-vein injection immediately after SCI and on postoperative days 1 and 2 | 21 days | BBB recovery, Evans blue leakage, lesion area, BSCB permeability, tight-junction proteins and endothelin-1-related rescue assessment |
| Morishima 2024 | Japan | Preclinical in vivo rat study with ex vivo/in vitro neutrophil assays | Female Sprague-Dawley rats; T6–T7 clip compression SCI | Amnion-derived MSC exosomes; miR-125a-3p-related mechanism | Tail-vein injection starting 24 h after SCI; three consecutive daily injections | 28 days | BBB recovery, lesion volume, neutrophil extracellular trap formation, MPO infiltration, inflammatory response and exosomal miR-125a-3p mechanism |
| Ran 2023 | China | Preclinical in vitro and in vivo mouse study | Female C57BL/6 mice; spinal cord contusion and crush models | Autologous plasma exosomes engineered with RVG, ILP and ISP peptides | Tail-vein injection starting 24 h after SCI; weekly repeated dosing | 6 weeks | BMS recovery, CatWalk gait parameters, MEP recovery, lesion repair, axonal regeneration, scar modulation, biodistribution and translational safety arm |
| Rao 2024 | China | Preclinical in vivo rat study with in vitro mechanistic assays | Male Sprague-Dawley rats; T10 impact contusive SCI | Hypoxic-preconditioned bone marrow MSC-derived small EVs | Caudal-vein injection 2 h after SCI and every 24 h for three total injections | 28 days | BBB recovery, MEP and CatWalk outcomes, lesion pathology, oxidative stress, apoptosis, neuronal survival and SIRT1/Nrf2/HO-1 pathway activation |
| Ruppert 2018 | USA | Preclinical in vivo rat study with EV characterization and immune profiling | Male Sprague-Dawley rats; T10 moderate contusive SCI | Human MSC-derived EVs, including EVs from inflammatory cytokine-primed MSCs | Single intravenous tail-vein injection 3 h after SCI | 14 days | BBB recovery, mechanical sensitivity, microglial/macrophage response, immune-cell profiling, lesion histology and inflammatory modulation |
| Wang 2021 | China | Preclinical biomaterial/EV in vitro and in vivo rat study | Female rats; complete T10 transection | Adipose MSC-derived EVs incorporated into injectable self-healing FE hydrogel | Orthotopic local injection into injured spinal cord immediately after transection | 56 days | BBB recovery, footprint outcomes, lesion repair, EV retention, glial scar response, inflammation, remyelination, axonal regeneration and hydrogel safety |
| Wang 2024 | China | Preclinical biomaterial/exosome in vitro and in vivo rat study | Female Sprague-Dawley rats; T9–T10 contusive SCI | Human iPSC-derived cortical neuron exosomes embedded in hUCMSC-derived decellularized ECM hydrogel | Local application/injection to lesion site during acute SCI surgery | 8 weeks | BBB recovery, inclined-plane and footprint outcomes, MEP recovery, tissue repair, bladder outcomes, angiogenesis, neural regeneration and hydrogel biocompatibility |
| Xiao 2021 | China | Preclinical in vivo rat study with mechanistic assays | Male Sprague-Dawley rats; T10 aneurysm-clip compression SCI | Human umbilical cord MSC-derived EVs; miR-29b-3p-depleted EV mechanistic arm | Tail-vein injection beginning 24 h after SCI; weekly repeated dosing | 28 days | BBB recovery, histopathology, Nissl staining, apoptosis, inflammatory markers, PTEN/Akt/mTOR signaling and miR-29b-3p mechanism |
| Zhang 2020 | China | Preclinical in vitro angiogenesis and in vivo mouse study | Male mice; T10 contusive SCI | Human placenta MSC-derived exosomes | Direct injection at SCI epicenter immediately after surgery | 42 days | BMS recovery, mechanical sensitivity, angiogenesis, micro-CT vascular analysis, endothelial tube formation, apoptosis and ANGPTL3-related pro-angiogenic effects |
| Zhou 2021 | China / Macau SAR | Preclinical in vivo rat transection study with in vitro NSC assays | Female Sprague-Dawley rats; complete thoracic transection at T11 | Human placental MSC-derived exosomes | Tail-vein injection 1 h after surgery and again at 2 weeks | 60 days | BBB recovery, bladder function, neurogenesis, NSC proliferation/differentiation, axonal/neural markers and endogenous repair activation |

Summary of the 24 included reports evaluating exosome- or extracellular vesicle-based interventions for spinal cord injury, including 23 animal/preclinical studies and one human/quasi-experimental study. The table includes study identification, publication year, country, study type, animal or participant characteristics where applicable, spinal cord injury model or clinical injury context, intervention source, delivery route, timing, follow-up duration, and main reported outcomes. The full extraction dataset is provided in Supplementary Table S2.

**Table 2. Risk-of-bias assessment of included studies.**

| No. | Study | Sequence generation | Baseline similarity | Random housing | Blinding of caregivers/investigators | Random outcome assessment | Blinding of outcome assessors | Incomplete outcome data | Selective outcome reporting | Other sources of Bias | Overall ROB |
| --- | --- | --- | --- | --- | --- | --- | --- | --- | --- | --- | --- |
| 1 | Cheng et al. 2026 | Low | Low | Unclear | Unclear | Low | Low | Unclear | Unclear | Unclear | Unclear |
| 2 | Su et al. 2026 | Low | Low | Unclear | Unclear | Low | Low | Unclear | Unclear | Unclear | Unclear |
| 3 | Li et al. 2026 | High | Unclear | Unclear | Unclear | Low | Unclear | Unclear | Unclear | Unclear | High |
| 4 | Li et al. 2025 | Unclear | Low | Unclear | Unclear | Low | Unclear | Unclear | Unclear | Unclear | Unclear |
| 5 | Lin et al. 2025 | Low | Low | Unclear | Unclear | Low | Low | Unclear | Unclear | Unclear | Unclear |
| 6 | Luo et al. 2025 | Unclear | Low | Unclear | Unclear | Low | Low | Unclear | Unclear | Unclear | Unclear |
| 7 | Deng et al. 2025 | High | Unclear | Unclear | Unclear | Low | Unclear | Unclear | Unclear | Unclear | High |
| 8 | Abbas et al. 2025 | High | Unclear | Unclear | Unclear | Low | Low | Unclear | Unclear | Unclear | High |
| 9 | Akhlaghpasand et al. 2024 | High | Unclear | Not applicable | Not applicable | High | High | Low | Unclear | Unclear | High |
| 10 | Zheng et al. 2024 | Low | Low | Unclear | Unclear | Low | Low | Unclear | Unclear | Unclear | Unclear |
| 11 | Morishima et al. 2024 | Unclear | Low | Unclear | Unclear | Low | Low | Unclear | Unclear | Unclear | Unclear |
| 12 | Rao et al. 2024 | Low | Low | Unclear | Unclear | Low | Low | Unclear | Unclear | Unclear | Unclear |
| 13 | Wang et al. 2024 | Low | Low | Unclear | Unclear | Low | Low | Unclear | Unclear | Unclear | Unclear |
| 14 | Akbari-Gharalari et al. 2023 | Unclear | Low | Unclear | Unclear | Low | Low | Unclear | Unclear | Unclear | Unclear |
| 15 | Ran et al. 2023 | Low | Low | Unclear | Unclear | Low | Low | Low | Unclear | Unclear | Unclear |
| 16 | Gao et al. 2021 | Low | Low | Unclear | Unclear | Low | Low | Unclear | Unclear | Unclear | Unclear |
| 17 | Wang et al. 2021 | Low | Low | Unclear | Unclear | Low | Low | Unclear | Unclear | Unclear | Unclear |
| 18 | Xiao et al. 2021 | Low | Low | Unclear | Unclear | Low | Low | Unclear | Unclear | Unclear | Unclear |
| 19 | Zhou et al. 2021 | Low | Low | Unclear | Unclear | Low | Unclear | Unclear | Unclear | Unclear | Unclear |
| 20 | Li et al. 2020 | Low | Low | Unclear | Unclear | Low | Unclear | Unclear | Unclear | Unclear | Unclear |
| 21 | Zhang et al. 2020 | Low | Low | Unclear | Unclear | Low | Low | Unclear | Unclear | Unclear | Unclear |
| 22 | Kang et al. 2019 | Low | Low | Unclear | Unclear | Low | Unclear | Unclear | Unclear | Unclear | Unclear |
| 23 | Guo et al. 2019 | Unclear | Low | Unclear | Unclear | Low | Low | Unclear | Unclear | Unclear | Unclear |
| 24 | Ruppert et al. 2018 | Low | Low | Unclear | Unclear | Low | Low | Unclear | Unclear | Unclear | Unclear |

Study-level risk-of-bias judgments using SYRCLE’s risk-of-bias tool for animal intervention studies. Domains include sequence generation, baseline similarity, random housing, blinding of caregivers/investigators, random outcome assessment, blinding of outcome assessors, incomplete outcome data, selective outcome reporting, and other sources of bias. One clinically oriented/quasi-experimental study was assessed using the JBI Critical Appraisal Checklist and is marked accordingly. Overall risk-of-bias categories were used descriptively and were not treated as weighted numerical scores.

**Table 3. Exosome source, engineering strategy, dose, route, and timing.**

| Study | Exosome / EV source | Engineering or delivery platform | Isolation / acquisition method | Dose / amount reported | Route | Administration timing and frequency | Follow-up |
| --- | --- | --- | --- | --- | --- | --- | --- |
| Su et al. 2026 | Regulatory T-cell-derived exosomes | Dual-engineered Treg-Exosome-IKVAV nanovesicles with IKVAV surface modification | Differential ultracentrifugation, iodixanol density gradient purification and dialysis | 2.0 × 10^11 particles/mL; 50 µL | Tail-vein intravenous injection | Single injection immediately after SCI | 28 days |
| Abbas et al. 2024 | iPSC-derived exosomes | Bioengineered P2X3-targeting exosomes with miRNA loading | Ultracentrifugation and/or exosome isolation reagent reported | 30 µg for biodistribution; therapeutic dose NR | Intravenous injection | Timing and number of therapeutic injections NR | 28 days |
| Akbari-Gharalari et al. 2023 | Platelet-rich plasma-derived exosomes | Dexamethasone-loaded PRP exosomes | Ultracentrifugation from activated human PRP | 10 µg exosomes in 100 µL PBS; 0.4 mg/kg; ExoDex every 3 days | Tail-vein intravenous injection | Started 1 h after SCI; repeated every 3 days | 28 days |
| Akhlaghpasand et al. 2024 | Human umbilical cord MSC-derived exosomes | Native allogeneic hUCMSC exosomes | Differential centrifugation, filtration and ultracentrifugation | 300 µg total exosomal protein in 5 mL; 60 µg/mL | Intrathecal injection via lumbar puncture | Single injection during subacute SCI phase, 1–5.5 months after injury | 12 months |
| Cheng et al. 2026 | hUCMSC-derived exosomes | Native hUCMSC exosomes combined with ROS-responsive trifluoperazine nanoparticles | Differential centrifugation and ultracentrifugation | Exosomes 2 mg/kg once weekly; nanoparticle dose 1 mg/kg | Systemic administration; imaging route intravenous | After SCI; exosomes once weekly for three injections | 6 weeks |
| Deng et al. 2026 | CD133+CD271+ human urine-derived stem-cell exosomes | Exosomes incorporated into injectable photosensitive CMCSMA hydrogel | Differential ultracentrifugation | 10 µL local hydrogel formulation; exosome amount NR | Local lesion-site injection with photocrosslinking | Single administration immediately after T10 transection | 56 days |
| Li et al. 2026 | HEK293T-derived engineered exosomes | circBDNF-loaded exosomes generated by donor-cell transfection | Differential ultracentrifugation | 10 µL exosome-derived circBDNF; particle/protein dose NR | Intrathecal injection at injury site | Immediately after SCI, then days 1 and 3 | 16 weeks |
| Gao et al. 2021 | M2 macrophage-derived exosomes | Berberine-loaded M2 macrophage exosomes | Ultracentrifugation | Exos-Ber 5 mg/kg; PBS 100 µL | Tail-vein intravenous injection | First dose 2 h after SCI; daily for 7 days | 28 days |
| Guo et al. 2019 | Human bone marrow MSC-derived exosomes | PTEN siRNA-loaded MSC exosomes | Differential centrifugation and ultracentrifugation | 40 µL per intranasal dose; 0.1 nmol PTEN-siRNA loaded into exosomes | Intranasal repeated dosing; intralesional comparator | Began 2–3 h after SCI; repeated daily for 5 days | 8 weeks |
| Kang et al. 2019 | MSC-derived exosomes | miR-21-enriched or PTEN-siRNA-related MSC exosomes | Commercial exosome isolation kit with high-speed/ultracentrifugation steps | Dose NR | Intravenous injection | After SCI; exact timing and number of injections NR | 28 days |
| Li et al. 2020 | Human MSC-derived exosomes | Exosomes immobilized in peptide-modified adhesive hydrogel scaffold | Size-exclusion chromatography followed by ultracentrifugation | Exo-pGel implanted into 4 ± 0.5 mm lesion gap; exosome amount NR | Local implantation into lesion gap; IV comparator | Single implantation immediately after long-span transection | 28 days |
| Li et al. 2025 | Umbilical cord MSC-derived exosomes | ucMSC exosomes combined with NSC-laden microfibers and hydrogel/scaffold system | Purchased ucMSC exosomes; supplier isolation method NR | 1 × 10^8/mL in vitro; in vivo dose NR | Local transplantation into complete spinal cord defect | Single intraoperative transplantation during acute SCI surgery | 4–8 weeks |
| Lin et al. 2025 | Adipose MSC-derived exosomes | Exosomes embedded in ROS-responsive injectable hydrogel | ADSC-exosome extraction reported; detailed protocol NR | 60 µL GEL-EXO injected locally; exosome amount NR | Local injection into transection site | Single injection immediately after complete transection | 6 weeks |
| Luo et al. 2026 | Bone marrow MSC-derived exosomes | Native BMSC exosomes combined with sodium tanshinone IIA sulfonate | Total exosome isolation kit | Exosomes 200 µg in 200 µL PBS; STS 10 mg/kg | Tail-vein intravenous injection plus oral gavage | Exosome single immediate injection; STS daily gavage | 21 days |
| Zheng et al. 2024 | Human umbilical cord MSC-derived exosomes | Native hucMSC exosomes | Ultracentrifugation | 200 µL at 200 µg/mL; approximately 40 µg per injection | Tail-vein intravenous injection | Immediately after SCI and postoperative days 1 and 2 | 21 days |
| Morishima et al. 2024 | Amnion-derived MSC exosomes | Native AMSC exosomes; miR-125a-3p investigated mechanistically | Differential centrifugation and ultracentrifugation | 100 µg exosomes in 1 mL PBS per injection | Tail-vein intravenous injection | First dose 24 h after SCI; three consecutive daily injections | 28 days |
| Ran et al. 2023 | Autologous plasma exosomes | AP-EXO-RVG/ILP/ISP peptide-decorated exosomes using CP05 anchor peptide | Differential centrifugation, filtration and ultracentrifugation | 1 mg/kg exosomes in 200 µL PBS | Tail-vein intravenous injection | Started 24 h after SCI; weekly for 4 weeks | 6 weeks |
| Rao et al. 2024 | Hypoxic-preconditioned bone marrow MSC small EVs | Hypoxia-preconditioned MSC-sEVs; no direct cargo/surface engineering | Density-gradient ultracentrifugation with sucrose layer | 500 µL at 0.2 µg/µL; 100 µg protein per injection | Caudal-vein intravenous injection | First injection 2 h after SCI, then every 24 h; three injections total | 28 days |
| Ruppert et al. 2018 | Human MSC-derived EVs | EVs from native or inflammatory cytokine-primed MSCs | Tangential flow filtration | 1 × 10^9 EVs/mL in 1 mL | Intravenous tail-vein injection | Single injection 3 h after SCI | 14 days |
| Wang et al. 2021 | Adipose MSC-derived EVs | EVs incorporated into injectable self-healing FE hydrogel | EV isolation reported; detailed method not visible in parsed main text | 10 µL local injection; therapeutic EV amount NR | Local orthotopic injection into injured spinal cord | Single injection immediately after complete transection | 56 days |
| Wang et al. 2024 | Human iPSC-derived cortical neuron exosomes | Exosomes embedded in hUCMSC-derived decellularized ECM hydrogel | Sequential centrifugation, Amicon concentration, precipitation reagent and ultracentrifugation | In vivo dose/injection volume NR | Local lesion-site application/injection | Single local treatment during acute SCI surgery | 8 weeks |
| Xiao et al. 2021 | Human umbilical cord MSC-derived EVs | Native HucMSC-EVs; miR-29b-3p-depleted EVs in mechanistic arm | Differential centrifugation and ultracentrifugation | 100 mg EVs as reported; injection volume NR | Tail-vein intravenous injection | First treatment 24 h after SCI; weekly injections until day 28 | 28 days |
| Zhang et al. 2020 | Human placental MSC-derived exosomes | Native hPMSC exosomes | Sequential centrifugation from hPMSC supernatant | 100 µg/mL in vitro; 200 µg/µL reported for in vivo use | Direct injection into SCI epicenter; intrathecal route also described | Single injection immediately after surgery | 42 days |
| Zhou et al. 2021 | Human placental MSC-derived exosomes | Native hpMSC exosomes | Ultracentrifugation; PEG precipitation compared but not used for main experiments | 50 µg total exosome protein in 100 µL PBS per injection | Tail-vein intravenous injection | At 1 h after surgery and again at 2 weeks; two injections total | 60 days |

Summary of intervention characteristics across included studies, including exosome or extracellular vesicle source, parent-cell origin, isolation method, engineering or cargo modification, biomaterial incorporation, dose reporting, administration route, number of injections, treatment timing after injury, and follow-up duration. This table highlights heterogeneity in exosome production, dosing, and delivery strategies.

**Table 4. Outcome domains and synthesis approach.**

| Outcome domain | Outcome status | Typical measures / markers | Data format extracted | Quantitative synthesis feasibility | Synthesis approach in this review |
| --- | --- | --- | --- | --- | --- |
| Locomotor functional recovery | Primary outcome | BBB score in rat studies; BMS score in mouse studies; gait or footprint-based motor assessments where reported | Mean, SD/SE, sample size, group, comparator, and follow-up timepoint | Partly feasible; exploratory BBB meta-analysis was possible in two studies with complete numerical data | Exploratory random-effects meta-analysis for BBB recovery where complete data were available; otherwise narrative synthesis |
| Lesion/cavity size and tissue preservation | Secondary outcome | Lesion area or volume, cavity size, spared tissue area, histological tissue preservation | Continuous data; mean, SD/SE, sample size, unit, and timepoint when available | Limited; complete comparable data were sparse | Descriptive synthesis; single-study standardized effects displayed where extractable |
| Neuronal survival and axonal/neural regeneration | Secondary outcome | NeuN, NF200, MAP2, TUJ1, GAP43, synaptophysin, axonal density, neurite growth, neural marker expression | Continuous marker expression, immunostaining intensity, positive-cell counts, or histological quantification | Limited; markers and measurement methods were heterogeneous | Descriptive synthesis grouped by regenerative outcome domain; single-study effects shown where possible |
| Myelination and oligodendrocyte preservation | Secondary outcome | MBP, myelin preservation, oligodendrocyte-related markers, remyelination indices | Continuous marker expression, staining intensity, positive-cell counts, or histological quantification | Not suitable for pooled meta-analysis because of heterogeneous markers and incomplete numerical reporting | Narrative synthesis emphasizing direction of effect and reported biological consistency |
| Glial scar and astrocytic response | Secondary outcome | GFAP expression, astrocyte activation, glial scar size or density, astrocyte phenotype-related markers | Continuous marker expression, staining intensity, or histological quantification | Not suitable for pooled meta-analysis | Descriptive synthesis by study and intervention type |
| Inflammation and immune modulation | Secondary outcome | TNF-α, IL-1β, IL-6, IL-10, CD68, Iba1, macrophage/microglial polarization, neutrophil infiltration, NET formation | Continuous protein/gene expression, immunostaining, cell counts, or cytokine concentrations | Not suitable for pooled meta-analysis because of heterogeneous markers and reporting formats | Narrative synthesis grouped by anti-inflammatory or immunomodulatory effect |
| Apoptosis and cell-death pathways | Secondary outcome | TUNEL staining, Bax, Bcl-2, cleaved caspase-3, apoptosis-related protein or gene expression | Continuous marker expression, staining intensity, positive-cell counts, or ratios | Limited; no formal pooled synthesis performed | Descriptive synthesis; effect direction reversed conceptually where lower values indicated benefit |
| Angiogenesis and vascular repair | Secondary outcome | CD31, VEGF, vascular density, micro-CT vascular metrics, endothelial markers, tube formation assays | Continuous vascular marker expression, vessel counts, vascular density, or imaging-derived measures | Limited; outcomes were heterogeneous and often study-specific | Descriptive synthesis; single-study effects displayed where complete data were available |
| Blood–spinal cord barrier integrity | Secondary outcome | Evans blue leakage, ZO-1, occludin, β-catenin, tight-junction proteins, permeability indices | Continuous permeability measures, protein expression, staining intensity, or leakage quantification | Limited; no formal pooled synthesis performed | Descriptive synthesis; lower-is-better outcomes interpreted as favorable when reduced by treatment |
| Neurogenesis and endogenous repair | Secondary outcome | NSC proliferation/differentiation markers, neural precursor markers, endogenous neurogenesis-related indices | Continuous marker expression, cell counts, or immunostaining quantification | Limited; mainly single-study or correlated marker data | Single-study standardized effects where extractable; otherwise narrative synthesis |
| Electrophysiological recovery | Secondary outcome | MEP amplitude, MEP latency, electrophysiological conduction recovery | Continuous amplitude or latency measures | Limited; insufficient comparable studies for pooling | Descriptive synthesis; single-study standardized effects where complete data were available |
| Autonomic and bladder-related recovery | Secondary outcome | Bladder function, urinary reflex recovery, residual urine, bowel/autonomic function where reported | Continuous or categorical functional data depending on study reporting | Not suitable for pooled meta-analysis | Narrative synthesis because measures were inconsistently reported |
| Safety and tolerability | Secondary outcome | Mortality, adverse events, body weight, systemic toxicity, organ histology, biodistribution, MRI safety, clinical safety outcomes | Event counts, proportions, continuous safety markers, or descriptive safety findings | Not suitable for pooled meta-analysis because reporting was inconsistent | Descriptive safety synthesis; clinical/quasi-experimental safety outcomes summarized separately |

Summary of primary and secondary outcomes reported across included studies and the corresponding synthesis method used in this review. Locomotor recovery was considered the primary outcome and was assessed using BBB or BMS scores. Secondary domains included lesion/cavity/tissue preservation, neuronal survival and axonal regeneration, myelination, glial scar response, inflammation, apoptosis, angiogenesis and blood–spinal cord barrier repair, neurogenesis, electrophysiology, and safety. Domains with complete numerical data were considered for quantitative synthesis; others were synthesized descriptively.

**Table 5. Exploratory quantitative synthesis of BBB locomotor recovery.**

| Study | SCI model | Intervention group | Comparator group | Timepoint | Treatment BBB score, mean ± SD | Control BBB score, mean ± SD | Effect measure | Effect estimate, Hedges’ g | Synthesis role | Interpretation |
| --- | --- | --- | --- | --- | --- | --- | --- | --- | --- | --- |
| Guo et al. 2019 | Complete T10 transection in rats | Intranasal MSC-derived exosomes loaded with PTEN siRNA | SCI transection control | 8 weeks | 7.75 ± 2.14; n = 7 | 0.39 ± 0.14; n = 15 | Standardized mean difference | 6.01; 95% CI 3.86 to 8.16 | Included in exploratory BBB meta-analysis | Favored exosome treatment |
| Li et al. 2020 | Long-span spinal cord transection in rats | hMSC-derived exosomes immobilized in adhesive hydrogel scaffold | Blank hydrogel/control lesion group | 28 days | 6.44 ± 1.64; n = 8 | 3.11 ± 2.13; n = 8 | Standardized mean difference | 1.66; 95% CI 0.48 to 2.83 | Included in exploratory BBB meta-analysis | Favored exosome treatment |
| Pooled estimate | Random-effects model | Exosome-based intervention | SCI control / comparator | Latest extractable follow-up | NA | NA | Standardized mean difference | 3.74; 95% CI −0.53 to 8.00 | Exploratory random-effects synthesis | Favored exosome treatment, but imprecise and crossed the null |

Study-level and pooled standardized mean differences for BBB locomotor recovery in studies with complete extractable mean, standard deviation, and sample-size data. Positive Hedges’ g values indicate greater locomotor recovery in exosome-treated animals compared with spinal cord injury controls. The pooled estimate was generated using a random-effects model and should be interpreted as exploratory because only two studies contributed data.

**Table 6. JBI critical appraisal of the included human phase I study.**

| **Study** | **Q1** | **Q2** | **Q3** | **Q4** | **Q5** | **Q6** | **Q7** | **Q8** | **Q9** | **Overall JBI appraisal** | **Risk-of-bias interpretation** |
| --- | --- | --- | --- | --- | --- | --- | --- | --- | --- | --- | --- |
| Akhlaghpasand et al. (2024) | Yes | Yes | Yes | No | No | Yes | Yes | Unclear | Unclear | Include | High risk for causal efficacy inference |

Q1, clear temporal relationship between cause and effect; Q2, participants included in comparisons were similar; Q3, participants received similar care apart from the intervention of interest; Q4, presence of an independent control group; Q5, multiple outcome measurements before and after the intervention; Q6, completeness of follow-up; Q7, outcomes measured consistently across comparisons; Q8, reliability of outcome measurement; Q9, appropriateness of statistical analysis. **Yes**, criterion satisfied; **No**, criterion not satisfied; **Unclear**, insufficient information for a definitive judgment. The nine questions correspond to the JBI Critical Appraisal Checklist for Quasi-Experimental Studies.

The study was retained because it provided directly relevant first-in-human safety and feasibility evidence. Its lack of an independent control group and multiple pre-intervention measurements substantially limited causal interpretation of efficacy outcomes. JBI does not prescribe a numerical summary score; the overall risk-of-bias interpretation was therefore assigned descriptively.
